# Transcutaneous Electrical Spinal Cord Stimulation Promotes Focal Sensorimotor Activation that Accelerates Brain-Computer Interface Skill Learning

**DOI:** 10.1101/2024.06.10.24308723

**Authors:** Hussein Alawieh, Deland Liu, Jonathan Madera, Satyam Kumar, Frigyes Samuel Racz, Ann Majewicz Fey, José del R. Millán

**Affiliations:** Chandra Family Department of Electrical and Computer Engineering, The University of Texas at Austin, Austin, USA; Department of Mechanical Engineering, The University of Texas at Austin, Austin, USA; Department of Neurology, The University of Texas at Austin, Austin, USA; Department of Biomedical Engineering, The University of Texas at Austin, Austin, USA

**Author notes:** **Correspondence** Correspondence should be addressed to: Hussein Alawieh and José del R. Millán.

## Abstract

Injuries affecting the central nervous system may disrupt neural pathways to muscles causing motor deficits. Yet the brain exhibits sensorimotor rhythms (SMRs) during movement intents, and brain-computer interfaces (BCIs) can decode SMRs to control assistive devices and promote functional recovery. However, non-invasive BCIs suffer from the instability of SMRs, requiring longitudinal training for users to learn proper SMR modulation. Here, we accelerate this skill learning process by applying cervical transcutaneous electrical spinal stimulation (TESS) to inhibit the motor cortex prior to longitudinal upper-limb BCI training. Results support a mechanistic role for cortical inhibition in significantly increasing focality and strength of SMRs leading to accelerated BCI control in healthy subjects and an individual with spinal cord injury. Improvements were observed following only two TESS sessions and were maintained for at least one week in users who could not otherwise achieve control. Our findings provide promising possibilities for advancing BCI-based motor rehabilitation.

## Introduction

Lesions in the corticospinal tract (CST) often cause long-lasting and severe motor impairments, carrying substantial personal and societal ramifications [1, 2]. Nevertheless, CST damage often spares functional neurons [3, 4] that brain-computer interfaces (BCIs) can draw on to decipher movement intentions from residual neural activity for the control of assistive devices [5–10]. Additionally, BCIs can engage users in neuromodulatory exercises with contingent feedback to elicit activity-dependent plasticity and facilitate recovery [11–16]. Such BCIs mainly rely on volitional modulation of sensorimotor rhythms (SMRs) via motor imagery (MI), the mental rehearsal of limb movement kinesthetics without physical execution [17, 18]. Electroencephalography (EEG) can capture SMRs non-invasively —revealing characteristic event-related desynchronization (ERD) and synchronization (ERS) within the *µ* (8-13 Hz) and *β* (13-30 Hz) frequency bands [18, 19]. These modulations are enhanced with motor learning [20], and the strength of ERDs within motor areas correlates with increased CST excitability [21–24].

Despite advancements in MI-based BCIs, a persistent challenge is the instability of SMR patterns due to the non-stationarity of EEG [25] that may significantly degrade BCI performance over days and hamper the effectiveness of BCI-based rehabilitation. Subjects often require longitudinal training to elicit reliable SMR and improve BCI accuracy [10, 26–32], so a fundamental question is how to accelerate subject training building upon the SMR neurophysiology. Previous attempts to enhance proficiency in SMR modulation either used rich haptic feedback or neuromodulatory techniques as an excitatory conditioning for the CST. Corbet et al. [33] illustrated how sensory-threshold neuromuscular electrical stimulation (St-NMES) contingent upon MI amplifies sensorimotor network activation and reinforces CST excitability. In [34], stroke patients undergoing NMES alongside BCI training exhibited stronger ERDs and increased laterality towards the ipsilesional motor area, which correlated with significant improvements in arm function. Similar results are observed in brain stimulation studies. Notably, *µ* ERDs showed significant increase following anodal transcranial direct current stimulation (tDCS), postulated to heighten cortical excitability by depolarizing neuron membrane potentials and increasing the firing likelihood during MI [35, 36]. While peripheral and brain stimulation methods have shown modulatory effects on SMRs, the potential of neuromodulation through electrical spinal stimulation remains untapped, despite its promising motor rehabilitation outcomes. Transcutaneous electrical spinal stimulation (TESS) increases the excitability of spinal motor pools, thus, increasing the likelihood that descending cortical drives reach target muscles [37]. Studies on TESS report increased muscle activations, heightened motor responses, and enhanced spinal reflexes during active movements [37–39]. TESS showcases therapeutic potential in restoring motor [37, 40–42] and autonomic functions [43, 44]. When examining the neuromodulatory effects of TESS, Benavides et al. [45] found that a single 20-minute TESS session delivered over C5/C6 vertebrae at 30 Hz with a 5 kHz carrier frequency (Fig. 1a) induced excitatory effects on the spinal level and increased cortical inhibition within the motor cortex. The latter manifested as increased short-interval intracortical inhibition (SICI) for up to 75 minutes post-stimulation.

**Figure 1:**
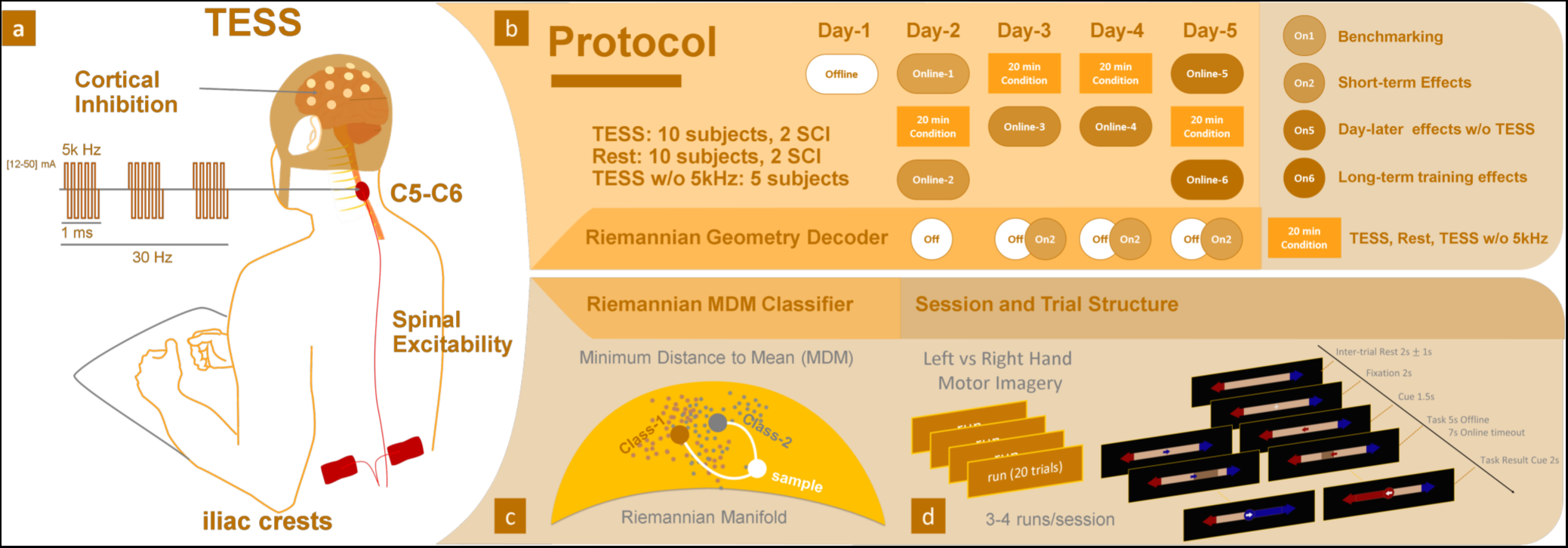
Experimental protocol. **a**: Transcutaneous Electrical Spinal Stimulation (TESS) was delivered at the cervical level between the C5-C6 spinal segment and the iliac crests of the hip bone using Hasomed’s RehaMove3 stimulator. We used blocks of five biphasic pulses delivered at 30 Hz with each pulse lasting for 200 us (carrier frequency of 5 kHz). **b**: the BCI training porotocol starts with an offline session on day-1 then a first baseline online session on day-2, which uses a classifier trained on the offline data. Starting on day-2, we begin applying the experimental condition (TESS) or the two control conditions (Rest or TESS w/o 5 kHz carrier) for 20 minutes before performing another online BCI session. Following day-2, the classifier is retrained using the data from the second online session. Days 3 and 4 start with 20 minutes of conditioning followed by an online BCI session. Day 5 starts with an online session to test one-day-later effects of training without further conditioning, then it proceeds with a final conditioning session followed by an online BCI session. **c**: Classification is performed using the illustrated minimum-distance-to-mean (MDM) Riemannian decoder, which uses covariance matrices of 22 EEG channels as features to classify left versus right motor imagery (MI). **d**: BCI sessions have 3-4 runs, each of which contains 20 trials split randomly and equally between left and right hand MI. The figure also illustrates the structure of a trial and the timings for each of its periods.

While the aim of most neuromodulation methods coupled to BCI has primarily been focused on increasing neural excitability to enhance SMR modulation —which, however, have not always led to better BCI performance [46–48]—, the potential role of inhibitory neuromodulation has been largely overlooked. Also, the role of inhibition in MI remains largely unknown. Recent evidence hints at elevated SICI during MI versus rest [49], implying that the excitatory motor output of MI might be counterbalanced by a parallel inhibitory output that prevents overt movement [49, 50]. While spinal-level inhibitory mechanisms are believed to be involved in motor preparation to safeguard motoneurons against premature activation, there is uncertainty about the origin of inhibitory mechanisms in MI [50]. Angelini et al. [51] argue that controlled inhibition of overt actions, as in Go/NoGo task, and automatic inhibition of covert actions, as in MI, share overlapping neural substrates. This can be the case in the “surround inhibition” phenomenon, which normally occurs in the motor cortex during overt movements to inhibit competing motor mechanisms allowing the execution of voluntary movements exclusively [52]. This phenomenon also appears in vivid MI [53] through which a focal ERD emerges surrounded by ERS.

These insights posit a role for inhibition in successful MI. Accordingly, we hypothesize that the inhibitory pre-conditioning of the cortex through TESS promotes focal SMR modulations —and, hence, reduces EEG non-stationarity—, resulting in more discriminable MI-related neural patterns, thus leading to faster MI skill acquisition and improved BCI control. In this work, we use a BCI based on motor imagery of hand movements, the usual approach in motor rehabilitation [12, 13, 21]. In particular, we chose right versus left hand MI that results in well-studied and reproducible SMR patterns with well-defined localization in the sensorimotor cortex [5, 19, 54] — allowing us to track the hypothesized increase in strength and focality of modulations with TESS. We validated the effectiveness of our intervention using a six-session longitudinal MI-based BCI training protocol (Fig. 1b), randomly assigning 20 healthy participants to either a test group subjected to 20 minutes of pre-training cervical TESS (TESS group) or a control group experiencing a 20-minute pre-training rest period (Rest group). Online session 2 of the protocol in Fig. 1b is intended to check the short-term effects of a single stimulation session while online session 5 tests the longer-term effects without further use of spinal stimulation. In addition to this core group of participants, we conducted complementary experiments to support the specific role of the inhibitory facet of TESS in promoting BCI skill learning. Finally,we explored the future potential of our intervention in motor rehabilitation by testing it with two SCI patients who completed the protocol twice, with and without spinal stimulation.

## Results

### Necessity of longitudinal SMR-based BCI training

The control of SMR-based BCIs typically requires feedback training over multiple sessions [10, 26–32]. This is evident in our study with the significant main effect of *session* in the analysis of variance for the linear mixed effects (LME-ANOVA) models that assess the results of both groups for all three BCI performance metrics (Fig. 2), namely Classification Accuracy (*F*_5_,_444_ = 4.85, *p <* 0.001); Bar Dynamics (*F*_5_,_444_ = 3.60*, p <* 0.01); and BCI Hits (*F*_5_,_444_ = 3.78, *p <* 0.01). Meanwhile, the percentage of timeouts across sessions for both groups showed no significant main effects (*Session* - 1 to 6: *F*_5_,_444_ = 1.48, *p* = 0.19, *Group* - TESS or Rest: *F*_5_,_444_ = 1.48, *p* = 0.19), nor significant *session* × *group* interaction effect (*F*_5_,_444_ = 0.68, *p* = 0.64). Specifically for the Rest group, a significant group-level increase in BCI performance —relative to the baseline in session 1— emerged in session 5 on the last day of training (Classification Accuracy: *t*_78_ = 3.90, *p_corrected_ <* 0.01; Bar Dynamics: *t*_78_ = 2.89, *p_corrected_ <* 0.05; and BCI Hits: *t*_77_ = 3.90, *p_corrected_ <* 0.05). Nevertheless, the Rest group still showed a significant increasing trend in performance across subjects for classification accuracy and BCI hits over the 6 training sessions (Fig 2b,c: *r >* 0.81*, p <* 0.05*, n* = 6) and a positive trend in bar dynamics (Fig 2a: *r* = 0.66*, p* = 0.096*, n* = 6). This is consistent with the expected learning process in longitudinal BCI training.

**Figure 2:**
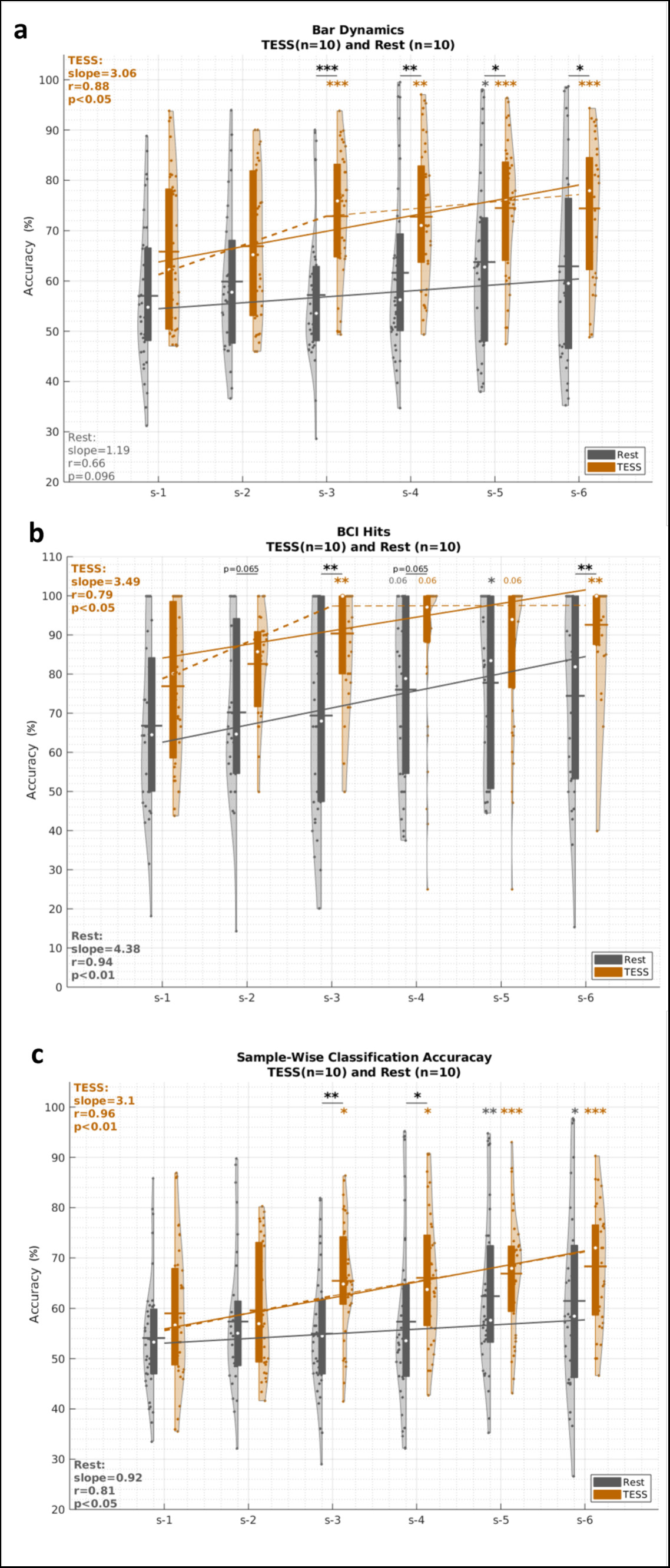
BCI performance results from the two main study groups. **a**: Bar dynamics, **b**: BCI hit accuracy, and **c** Sample-wise classification accuracy, for the TESS (in orange) and Rest (in gray) groups over the six BCI sessions of the protocol. Violin plots show values of the (subject,run) pairs. Boxes within violins correspond to the inter-quartile range, white circles correspond to medians, and horizontal lines correspond to means. Session 1 performance serves as a baseline for within-group comparisons. LME-ANOVA reveal a significant main effect of session forall three metrics (*F*_5_,_444_ ≥ 3.6*, p <* 0.01) and a significant *session* × *group* interaction (*F*_5_,_444_ = 2.06*, p <* 0.05) for BCI hits. Post-hoc paired tests showed significant improvement in all BCI performance metrics for the TESS group starting on session 3, after two stimulation sessions, that persisted till the end of the training while significance emerged only after session 5 for the Rest group. Between-subject post-hoc tests show that TESS significantly outperforms Rest in BCI control (bar dynamics) and command delivery (BCI hits) starting on session 3. The solid regression lines show the trends of the medians over sessions. The dashed lines correspond to piece-wise spline regression of medians. Both groups show significantly increasing trends over sessions. Significant differences across groups are indicated by underlined asterisk, and within-group significant difference relative to baseline is indicated above the violins with orange asterisks for TESS and gray asterisks for Rest. Statistical testing is performed using linear mixed effect models. ∗ : *p <* 0.05, ∗∗ : *p <* 0.01, ∗∗∗ : *p <* 0.001, absence of asterisks indicates non-significant difference. All *p*-values are Bonferroni-Holm corrected for multiple comparisons.

### TESS accelerates BCI training and MI skill learning

While both TESS and Rest groups showed a learning effect throughout training, the TESS group exhibited a significant enhancement of BCI performance relative to baseline starting as early as session 3 —compared to session 5 for the Rest group— (Classification Accuracy: *t*_78_ = 3.30, *p_corrected_ <* 0.05; Bar Dynamics: *Z* = 3.97, *p_corrected_ <* 0.001; and BCI Hits: *t*_75_ = 4.38, *p_corrected_ <* 0.01). Performance was further enhanced at the end of training on session 6 (Classification Accuracy: Δ*_CA_* = 9.31%, *t*_78_ = 4.98, *p_corrected_ <* 0.001; Bar Dynamics: Δ*_barDyn_* = 8.64%, *t*_73_ = 4.77, *p_corrected_ <* 0.001; and BCI Hits: Δ*_Hits_* = 15.71%, *t*_70_ = 4.58, *p_corrected_ <* 0.001). For both sessions 3 and 6, the TESS group showed a large effect size for the BCI hits improvement (Cohen’s *d_s_*_3_ = 0.81, *d_s_*_6_ = 0.94). The TESS group also maintained significant improvement in performance on session 5, which occurred at the start of a new training day and was not preceded by stimulation. TESS significantly outperformed Rest on session 3 for all performance metrics (*t*_78_ *>* 3.96, *p_corrected_ <* 0.01) and maintained significantly higher bar dynamics and BCI hits on the last session (*t*_66_ *>* 2.88, *p_corrected_ <* 0.01). Over the course of the sessions, the TESS group showed an increasing trend in BCI bar dynamics control (Fig 2a, *slope* = 3.06*, r* = 0.88*, p <* 0.05*, n* = 6). Since BCI hits saturated close to 100% following session 4 for TESS, we identified the presence of an initial steep learning slope reflecting rapid gains in the early learning phase followed by a gradual asymptotic plateau, consistent with the power law of practice [55]. The trend of the BCI hits accuracy in the TESS group from Session 1 to Session 3, the inflection point, was significantly increasing (*slope* = 9.29*, r* = 0.90*, p <* 0.05*, n* = 3).

### Strong and focal ***µ*** and ***β*** ERDs emerge with TESS

Having demonstrated how the proposed intervention facilitates participants’ acquisition of BCI control, we examine the neurophysiological underpinnings of such BCI skill learning process. To this end, we analyzed SMR modulations in EEG during training, in particular the *µ* and *β* band ERDs in the contralateral sensorimotor areas. For analysis purposes, right MI data were mirrored before being pooled with left MI data so that ERDs are localized on the right hemisphere. For each subject, we first identified the single EEG channel in the contralateral sensorimotor cortex that exhibited the largest ERD strength at the end of training (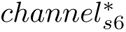) and backtracked its ERD over sessions. Fig. 3a,b show the *µ* ERD for 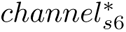 averaged across subjects in the TESS and Rest groups, respectively. In Fig. 3a, TESS exhibits significantly larger *µ* ERD in session 6 relative to session 1 (Cluster permutation testing: interval [3.53, 4.89] s, *p_cluster_* = 0.041 and interval [5.27, 8.5] s, *p_cluster_* = 0.017), in contrast to Rest. The inset topoplot for session 6 in TESS shows the emergence of a more focal ERD source at the relevant C3 location compared to that of Rest. Fig. 3c also illustrates that the contra-lateral ERD difference between session 6 and session 1 —averaged over the task execution period— is stronger for the TESS group relative to the Rest group. In the *β* band, both groups show significantly larger ERD in session 6 relative to session 1, but the difference is more prominent for TESS (TESS: [0.40, 5.11] s, *p* = 0.008; Rest: [1.63, 4.78] s, *p* = 0.012). When averaged over the task execution period, the ERDs in Fig. 3f show stronger change in contraletral ERD for TESS relative to Rest —consistent with the observation for the *µ* ERDs in Fig. 3c. To further analyze the focality of the emerging ERD patterns, we normalized the strength of the ERD from 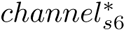 to the ERDs within the motor area using equation (6). We refer to the latter as *ERD*^∗^. Fig. 3g,h show the evolution of *ERD*^∗^ for the *µ* and *β* bands, respectively. LME-ANOVA revealed significant effect of *session* on *ERD*^∗^ in both *µ* (*F*_5_,_444_ = 3.19*, p* = 0.008) and *β* (*F*_5_,_444_ = 6.50*, p <* 0.001). Only the TESS group exhibited a significantly increasing focality of *µ* ERD —i.e., a decreasing trend of *ERD*^∗^ over the 6 sessions (*slope* = −0.08, *r* = −0.23, *p <* 0.001, *n* = 6) in Fig. 3g. On session 5, which was not preceded by stimulation, the *µ ERD*^∗^ for the TESS group became significantly larger than baseline (*t*_77_ = −3.05, *p_corrected_* = 0.015) and also significantly larger than that of the Rest group (*t*_77_ = −3.43, *p_corrected_* = 0.008). At the end of the training on session 6, both groups had significantly enhanced *ERD*^∗^ (Rest: *t*_71_ = −3.01, *p_corrected_* = 0.015; TESS: *t*_73_ = −5.18, *p_corrected_ <* 0.001), but TESS remained significantly larger than Rest (*U* = 2.59*, p* = 0.031, Mann-Whitney U test) showing a large effect size (Cohen’s *d* = 0.94) compared to a medium effect for Rest (Cohen’s *d* = 0.58). For the beta *ERD*^∗^ in Fig. 3h, both groups showed significantly decreasing trends (TESS: *slope* = −0.09, *r* = −0.28, *p <* 0.001, *n* = 6; Rest: *slope* = −0.08, *r* = −0.21, *p* = 0.002, *n* = 6). Participants in the TESS group, however, achieved significantly larger 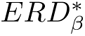 —relative to their baseline— on session 4 (*t*_78_ = −3.76, *p_corrected_* = 0.002) while the Rest group reached significance only on session 6 (Rest: *t*_71_ = −4.78, *p_corrected_ <* 0.001). Overall, ERD results support the role of TESS in promoting faster and more effective modulation of SMRs for healthy subjects.

**Figure 3:**
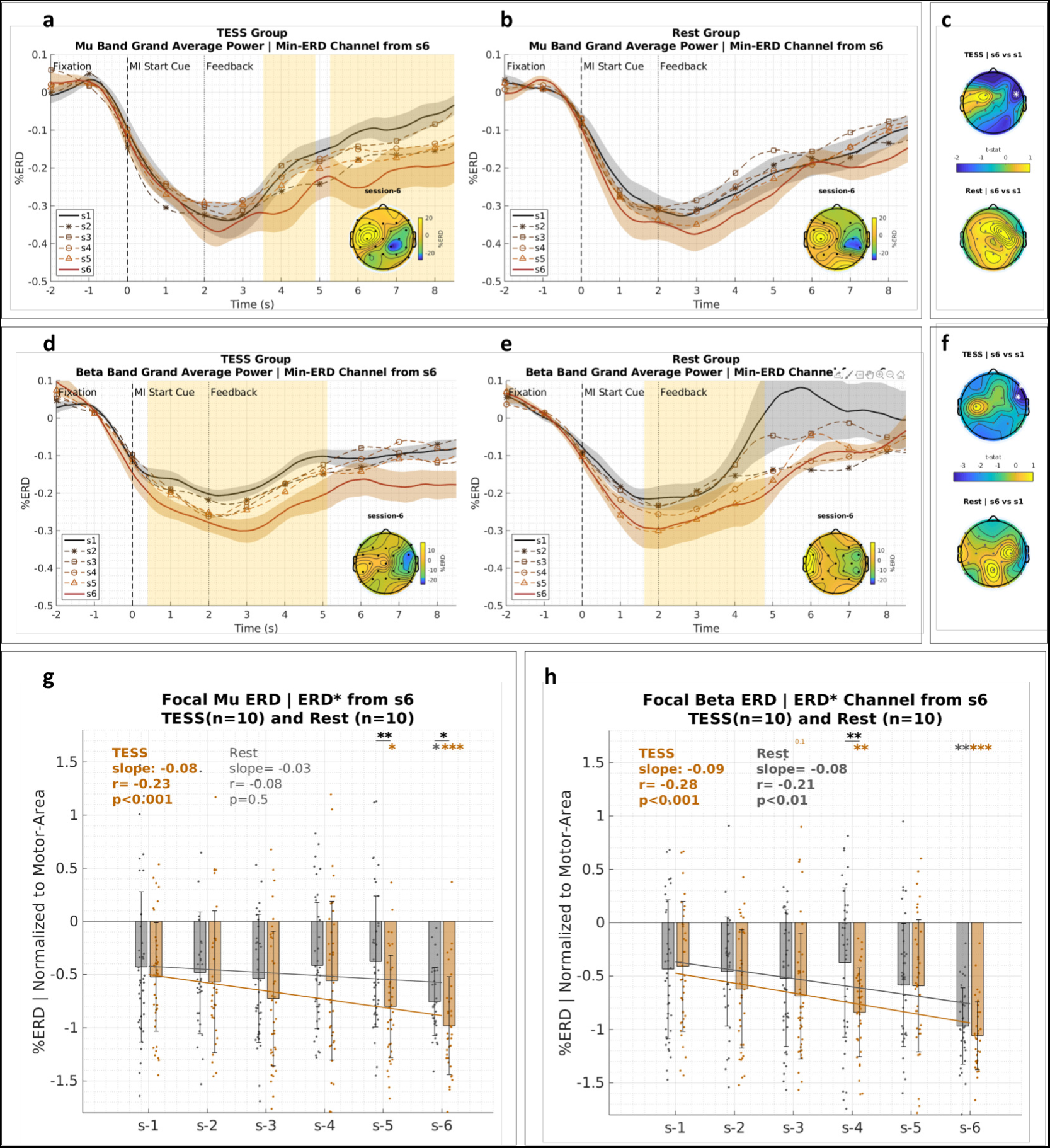
Neurophysiological evidence of enahnced SMR modulations. **a**, **b**: Grand average ERD patterns in the *µ* band for the channel that exhibited the highest ERD strength at the end of training in the TESS and Rest groups, respectively. Black and orange lines correspond to the first and last online sessions, respectively. Inset topoplots show the ERD patterns over selected EEG channels in session 6. The confidence levels for the grand average patterns represent standard deviations across subjects, and the shaded time periods represent statistically significant difference between session 1 and 6 with cluster-based permutation testing. **c**: t-statistic of the differences in ERD strength between the last and the first session for the TESS group on top and the Rest group at the bottom. White asterisks indicate statistically significant difference using cluster permutation testing. **d**, **e**, and **f** : ERD patterns and topography for the *β* band similar to *µ* band above. **g**, **h**: Focality of ERD patterns in the *µ* and *β* bands, respectively, for the TESS (orange) and Rest (gray) groups. The plots backtrack the ERD strength of the channel that exhibited the greatest desynchronization at the end of training (*ERD*^∗^) after normalizing it to the ERD values within the motor area (i.e., the FC, C, and CP electrodes). Trend lines are regression fits of session means. Significant differences across groups are indicated by underlined asterisk, and within-group significant difference relative to baseline is indicated above the bars with orange asterisks for TESS and gray asterisks for Rest. Statistical testing is performed using linear mixed effect models. ∗ : *p <* 0.05, ∗∗ : *p <* 0.01, ∗∗∗ : *p <* 0.001, absence of asterisks indicates non-significant difference. All p-values are Bonferroni-Holm corrected for multiple comparisons. Error bars represent standard deviations.

### TESS helps slow BCI learners gain control

We further challenged the ability of TESS to help slow BCI learners or non-responders to typical training paradigms. For that, we identified the individuals in the Rest group who could not learn to control the BCI after the six training sessions. A total of four participants had no control over the bar dynamics at the end of training (Supplementary Fig. 1). These subjects were re-enrolled for the TESS intervention after a 6 months washout period during which they were not involved in any BCI study. Additionally, they also completed a follow-up BCI session —without TESS— one week after they finished the TESS protocol to test for long-lasting effects. After training with TESS, all subjects had improved control. LME-ANOVA shows a significant *session* × *modality* interaction effect for bar dynamics (*F*_5_,_168_ = 2.48, *p* = 0.034) and BCI hits (*F*_5_,_168_ = 3.96, *p <* 0.01). Only when trained with TESS, these subjects could achieve significantly higher bar dynamics accuracy at the end of training on both session 5 (*t*_30_ = 4.44, *p_corrected_ <* 0.01), which was no longer preceded with stimulation, and session 6 (*t*_27_ = 4.55, *p_corrected_ <* 0.01) (Fig. 4a). This is also the case for the BCI hits metric (Fig. 4b; session 5: *t*_30_ = 4.95, *p_corrected_ <* 0.001; session 6: *t*_27_ = 5.19, *p_corrected_ <* 0.001). The subjects could actually achieve significantly better BCI hits accuracy with TESS starting on session 3 (*t*_30_ *>* 4.16, *p_corrected_ <* 0.01) with a very large effect size (Cohen’s *d >* 1.34) and they maintained it till the end of training. Overall, the subjects showed a significantly increasing trend in both metrics over the 6 sessions (Bar Dynamics: *r* = 0.94*, p <* 0.01*, n* = 6; BCI Hits: *r* = 0.96*, p <* 0.01*, n* = 6), and they could maintain significantly higher performance relative to their baseline on the one-week follow up session after TESS (Bar Dynamics: *t*_28_ = 3.77, *p_corrected_* = 0.015; BCI Hits: *t*_28_ = 3.89, *p_corrected_ <* 0.01) with a large effect size (Cohen’s *d >* 1.042). Fig. 4c backtracks 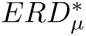 over the six training sessions of the protocol. Consistent with the BCI performance results, the subjects showed significantly increasing focality in 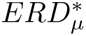 only when trained with TESS (*r* = −0.44*, p <* 0.001). The latter increase in 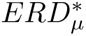 strength became significant at the end of training (*t*_28_ = −5.88, *p_corrected_ <* 0.001) with a very large effect size (Cohen’s *d* = 2.12), and it was already tending towards significance on session 5 (*t*_28_ = −2.71, *p_corrected_* = 0.058). Fig. 4d,e show the grand average *µ* ERD patterns for 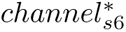 . Only TESS shows a period of significantly larger *µ* ERD on session 6 compared to session 1 (interval: [1.07, 5.68] s, *p_cluster_* = 0.028). Supplementary Fig, 2 illustrates the results for *β* band ERD. Both modalities exhibited an increasing *β* ERD focality over sessions (Supplementary Fig. 2c); TESS: *r* = −0.26*, p <* 0.05; Rest: *r* = −0.30*, p <* 0.01), consistent with the original group. The increase reached significant difference relative to baseline for the Rest modality on session 6 (*t*_27_ = −3.42*, p_corrected_* = 0.032); however, none of the modalities showed a significant change in the grand average ERD patterns (Supplementary Fig. 2a,b).

**Figure 4:**
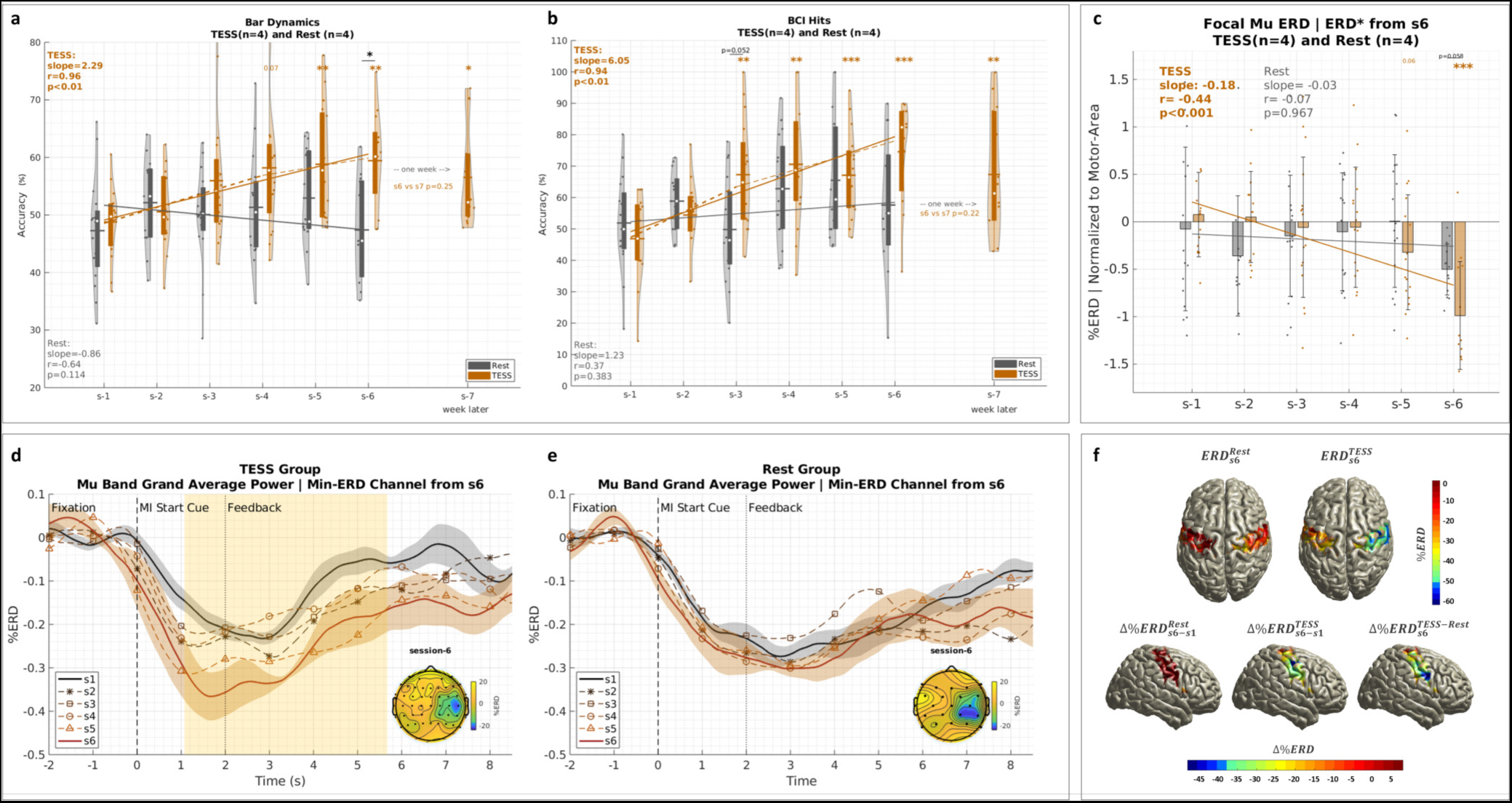
Cross-Over group of slow learners. **a** and **b**: Bar dynamics and BCI hits performance, respectively, similar to Fig. 2a,b for the set of slow BCI learners when they did the intervention without stimulation (gray) and when they repeated it six months later with TESS (orange). **c**: Focality of *µ* ERD patterns (*ERD*^∗^) for the TESS (orange) and Rest (gray) groups similar to Fig. 3g. **d**, **e**: Grand average *µ* ERD patterns for the channel that showed the highest ERD strength at the end of training similar to Fig. 3a, b for the TESS and Rest modalities, respectively. The shaded region in the TESS case highlights the period of significant difference between sessions 1 and 6. **f** : Source level analysis of *µ* ERDs. The top part shows session 6 patterns for the Rest (left) and TESS (right) modalities while the bottom part focuses on the contra-lateral hemisphere to show —from left to right—the difference between session 6 and session 1 for Rest and TESS as well as the difference at session 6 between the two modalities. ∗ : *p <* 0.05, ∗∗ : *p <* 0.01, ∗∗∗ : *p <* 0.001, absence of asterisks indicates non-significant difference. All p-values are Bonferroni-Holm corrected for multiple comparisons. Error bars represent standard deviations.

To further analyze the physiological differences that emerge when training with TESS, we used subject-specific MRIs to reconstruct the source activity at the cortical level. We focused on the *µ* band where a significant change in modulation was observed. Fig. 4f shows the average source patterns after warping the individual patterns of each subject on a common MNI template brain. The top part of Fig. 4f shows the *µ* ERD patterns in session 6, which were very weak on the contra-lateral right hemisphere when subjects trained without stimulation (left) compared to when they re-trained with TESS (right). The bottom part shows the *µ* ERD differences on the contra-lateral right hemisphere between session 6 and session 1 for each modality as well as the difference between modalities on the last session. Clearly, there is stronger desynchronization with TESS —consistent with the channel-level results in the left plot of Fig. 4d. The pattern differences on the last session also show higher *µ* ERD for TESS compared to Rest, and this appears close to where the hand representation is expected to be in the contra-lateral hemisphere. Per-session source results are provided in Supplementary Fig. 3, and they show evolving ERD patterns on the contra-lateral motor area consistent with the subjects’ incremental skill learning over sessions in Fig. 4a,b.

### Cortex-exciting spinal stimulation impedes BCI learning

Since our hypothesis advocates a role of cortex-inhibiting TESS in promoting BCI skill learning, we also investigated a potential opposite effect for cortex-exciting spinal stimulation. The latter can be achieved through a modified stimulation sequence that does not incorporate the 5 kHz carrier frequency as detailed and validated in [45] (refer to Fig. 6). Five additional subjects were recruited to complete the experimental protocol of Fig. 1b with excitatory spinal stimulation instead of TESS (“No-Carrier” group). Fig. 5a,b show the BCI performance of these subjects compared to the original TESS group. LME-ANOVA shows a significant *session* × *group* interaction effect (Bar Dynamics: *F*_5_,_328_ = 4.87*, p <* 0.001, BCI Hits: *F*_5_,_328_ = 2.73*, p* = 0.019) with the No-Carrier group showing no significant change over the six training sessions. In fact, statistical testing with the Rest group also shows a significant *group* × *session* interaction effect for both BCI performance metrics (*F*_5_,_324_ *>* 3.13*, p <* 0.01) suggesting that the subjects in the No-Carrier group were impeded from acquiring better BCI control. Fig. 5a even shows a significant decrease in bar dynamics accuracy for the No-Carrier group on the last day of training (session 5: *t*_38_ = −2.62*, p_corrected_* = 0.023). This is also consistent with the fact that the ERD patterns in Fig. 5c,d for the *µ* and *β* bands remained almost unchanged throughout the training. The inset topoplots show strong ERDs on session 6; however, they are less localized relative to those of the TESS group (Fig. 3a,d). Nevertheless, when normalized within the motor area, 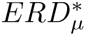 in Fig. 5c shows a marginally significant trend over sessions (*r* = −0.21*, p* = 0.052), but it was not as prominent as that of the TESS group (*r* = −0.28*, p <* 0.001). Meanwhile, *β* ERDs in Fig. 5f show no significant change. Results from this group provide further evidence that the cortex-inhibiting facet of TESS plays the major role in promoting SMR modulation and BCI skill learning.

**Figure 5:**
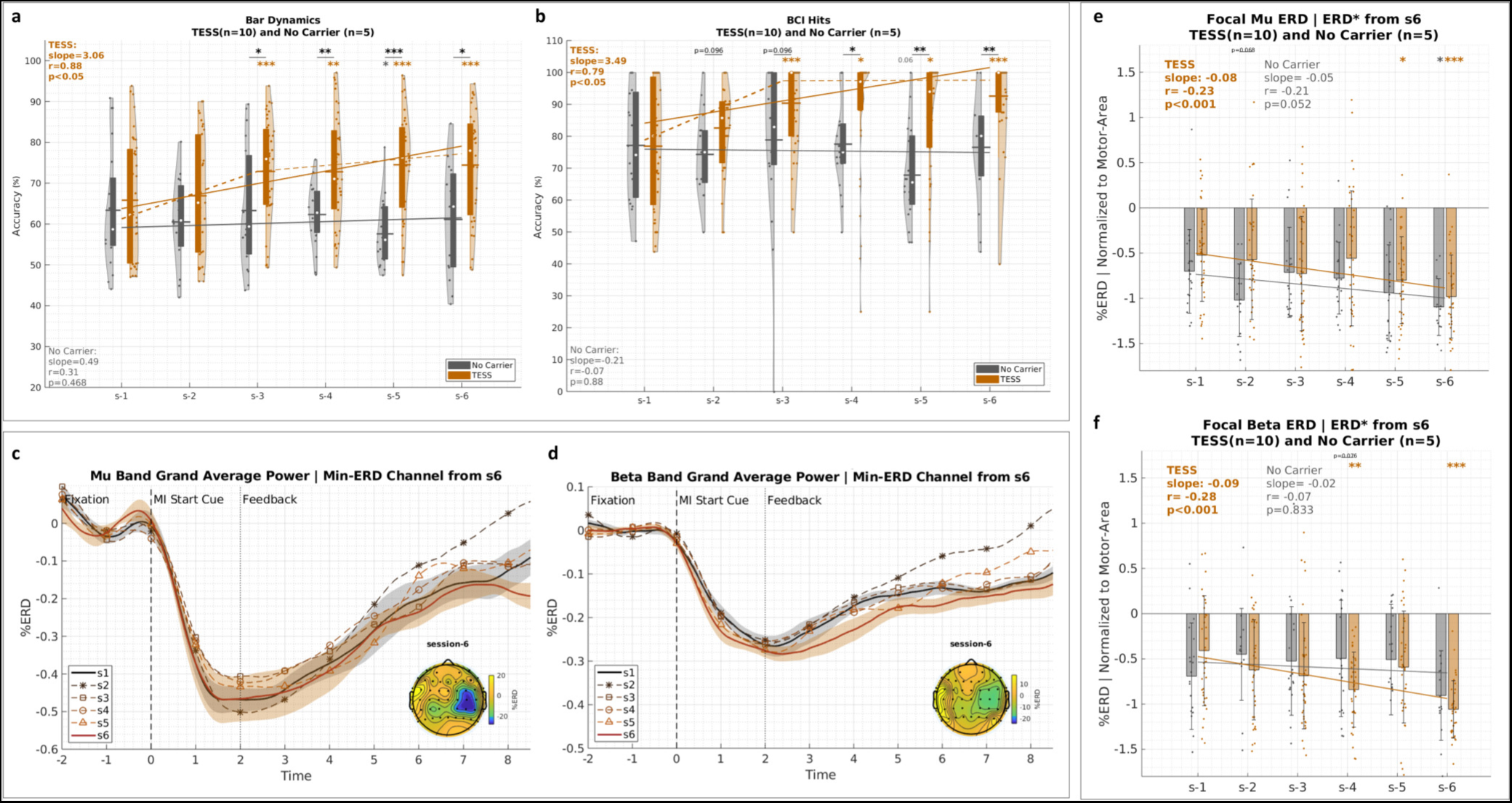
No-Carrier group. **a**, **b**: Bar dynamics and the BCI hits, respectively, for the No-Carrier group (gray) in comparison to the original TESS group (orange) similar to Fig. 2a,b. **c**, **d**: Grand average *µ* and *β* ERD patterns, respectively, for the channel that showed the highest ERD strength at the end of training similar to Fig. 3a, d. **e**, **f** : Focality of *µ* and *β* ERD patterns (*ERD*^∗^), respectively, for the TESS (orange) and Rest (gray) groups similar to Fig. 3g,h. ∗ : *p <* 0.05, ∗∗ : *p <* 0.01, ∗∗∗ : *p <* 0.001, absence of asterisks indicates non-significant difference. All p-values are Bonferroni-Holm corrected for multiple comparisons. Error bars represent standard deviations.

**Figure 6:**
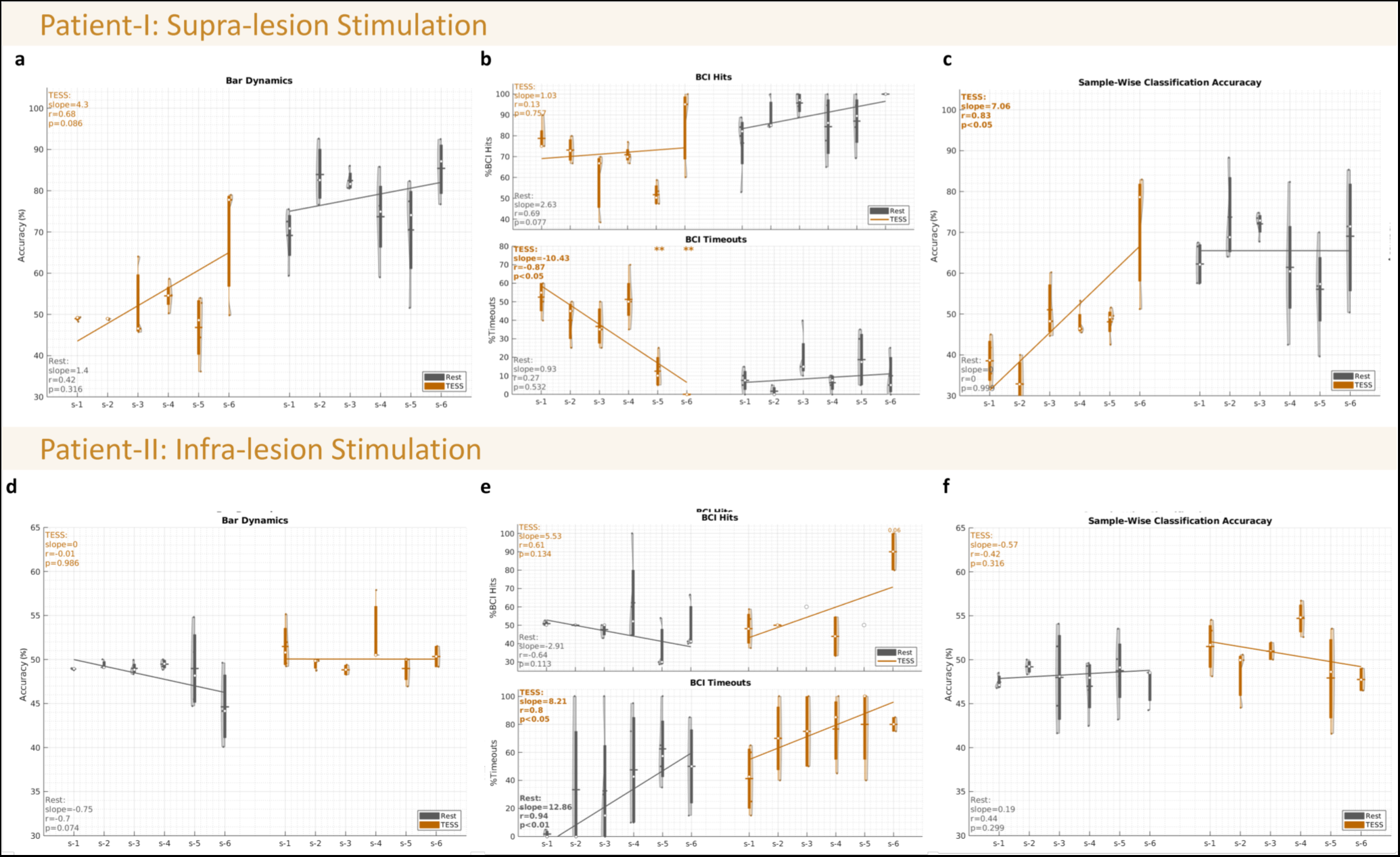
BCI performance results for the SCI patients. **a**, **b**, and **c**: Bar dynamics, BCI hit accuracy, and sample-wise classification accuracy, respectively, for Patient-I who received supra-lesional stimulation. **d**, **e**, and **f** : Bar dynamics, BCI hit accuracy, and sample-wise classification accuracy, respectively, for Patient-II who received infra-lesional stimulation. Violin plots show values of the (subject,run) pairs. Boxes within violins correspond to the inter-quartile range, white circles correspond to medians, and horizontal lines correspond to means. Session 1 performance serves as a baseline for within-group comparisons. The solid regression lines show the trends of the medians over sessions.

### Supra-lesional TESS enhances BCI learning post-SCI

To illustrate the potential of the proposed intervention in motor rehabilitation and the control of assistive devices, we enrolled two subjects with tetraplegia after SCI who completed the experimental protocols for TESS and Rest with one week of break. Patient-I had a C5 fracture and received supra-lesional stimulation while Patient-II had a C4 fracture and received infra-lesional stimulation. Results in Fig. 6a,c for Patient-I show a significant main effect for TESS (Classification Accuracy: *F*_1_,_31_ = 14.44, *p <* 0.001; Bar Dynamics: *F*_1_,_31_ = 14.87, *p <* 0.001) with a significant increase in classification accuracy (*r* = 0.83*, p* = 0.021) and an increasing trend in bar dynamics (*r* = 0.68*, p* = 0.086). This increase during the TESS intervention sessions was followed by a plateau, or a less steep slope, one week later when the patient resumed the study without TESS. Although the BCI hit accuracy in the upper part of Fig. 6b shows no significant change over the sessions throughout the entire experiment, the overall number of delivered commands increased during TESS for Patient-I because, as shown in the lower part of Fig. 6b, there was a significantly decreasing number of timeout trials (*r* = −0.86*, p* = 0.013). The latter exhibited a significant decrease on the last day of training (session 5: *t*_6_ = −6.82, *p_corrected_ <* 0.01; session 6: *t*_5_ = −10.97, *p_corrected_ <* 0.01). The results also show a significant *session* × *intervention* interaction effect for both classification accuracy (*F*_5_,_31_ = 4.34, *p <* 0.01) and timeouts ratio (*F*_5_,_31_ = 13.19, *p <* 0.001) supporting a period of skill learning and consolidation during TESS followed by a period of maintained performance one week later without TESS. When analyzing ERD patterns for the Patient-I, results reveal a significant *session* × *group* interaction effect for 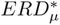 (*F*_5_,_31_ = 7.78*, p <* 0.001) and 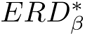 (*F*_5_,_31_ = 9.45*, p <* 0.001). The patient showed a prominent change in ERD patterns on session 3 (day 2), which coincides with the re-calibration of the decoder to incorporate emerging patterns following the first post-stimulation online session (session 2). Starting on session 3 onwards, both 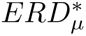 and 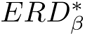 show significantly strong trends (*µ*: *slope* = −0.77*, r* = −0.78*, p <* 0.001; *β*: *slope* = −0.4*, r* = −0.78*, p <* 0.01) followed by a period of non-significant change one week later when the patient repeated the protocol without stimulation (Supplementary Fig. 4a,b).

Regarding Patient-II, as expected because of the infra-lesional stimulation, both Rest and TESS modalities did not result in a considerable change in performance (Fig. 6d,f). However, the upper part of Fig. 2e shows an increase in BCI hits during the TESS sessions (session 6: *t*_4_ = 5.39, *p_corrected_* = 0.057) with a significant *session* × *intervention* interaction effect (*F*_5_,_18_ = 4.03, *p* = 0.013). Nevertheless, this came at the expense of an increasing number of timeouts.

### Physiological correlates of enhanced BCI control following TESS

Finally, we performed additional analyses to better understand the neurophysiological basis of how TESS promotes BCI skill acquisition. Spearman’s correlation analysis revealed that the increase in *µ* ERD focality moderately explained better BCI command delivery performance in all the healthy users (Fig. 7a, Spearman correlation: *r* = −0.4*, p <* 0.001*, n* = 165) —i.e., including all 29 subjects in the TESS, Rest, Cross-Over, and No-Carrier groups. The correlation was also significant for the TESS subjects (Spearman correlation: *r* = −0.48*, p <* 0.001*, n* = 77). For Patient-I, who received supra-lesional stimulation and showed evidence of BCI learning, the correlation was particularly larger (Spearman correlation: *r* = −0.6*, p* = 0.028). We further checked the specific role of stimulation-induced inhibition in promoting ERD focality and BCI performance. For that we used resting-state EEG recordings —collected for the subjects in the Cross-Over and No-Carrier groups as well as Patient-I— to study the effect of stimulation on the *α* band power, which is commonly associated with inhibitory processes [56–58]. Fig. 7b shows that TESS results in a significant increase in resting-state *α* power within the motor cortex (Δ*ERS* = 26.75% ± 17.42%*, t*_4_ = 3.43*, p* = 0.026) while the no-carrier stimulation has no significant effect (Δ*ERS* = 5.67% ± 14.74%*, t*_4_ = 0.86*, p* = 0.439). The strength of *α* power post-stimulation explains the increased focality of *µ* ERD (Supplementary Fig. 5: Spearman correlation, *r* = −0.58*, p <* 0.001*, n* = 40), and Fig. 7c shows that the percentage change in *α* power post-stimulation correlates with the percentage change in classification accuracy (Spearman correlation: *r* = 0.41*, p <* 0.01*, n* = 40).

**Figure 7:**
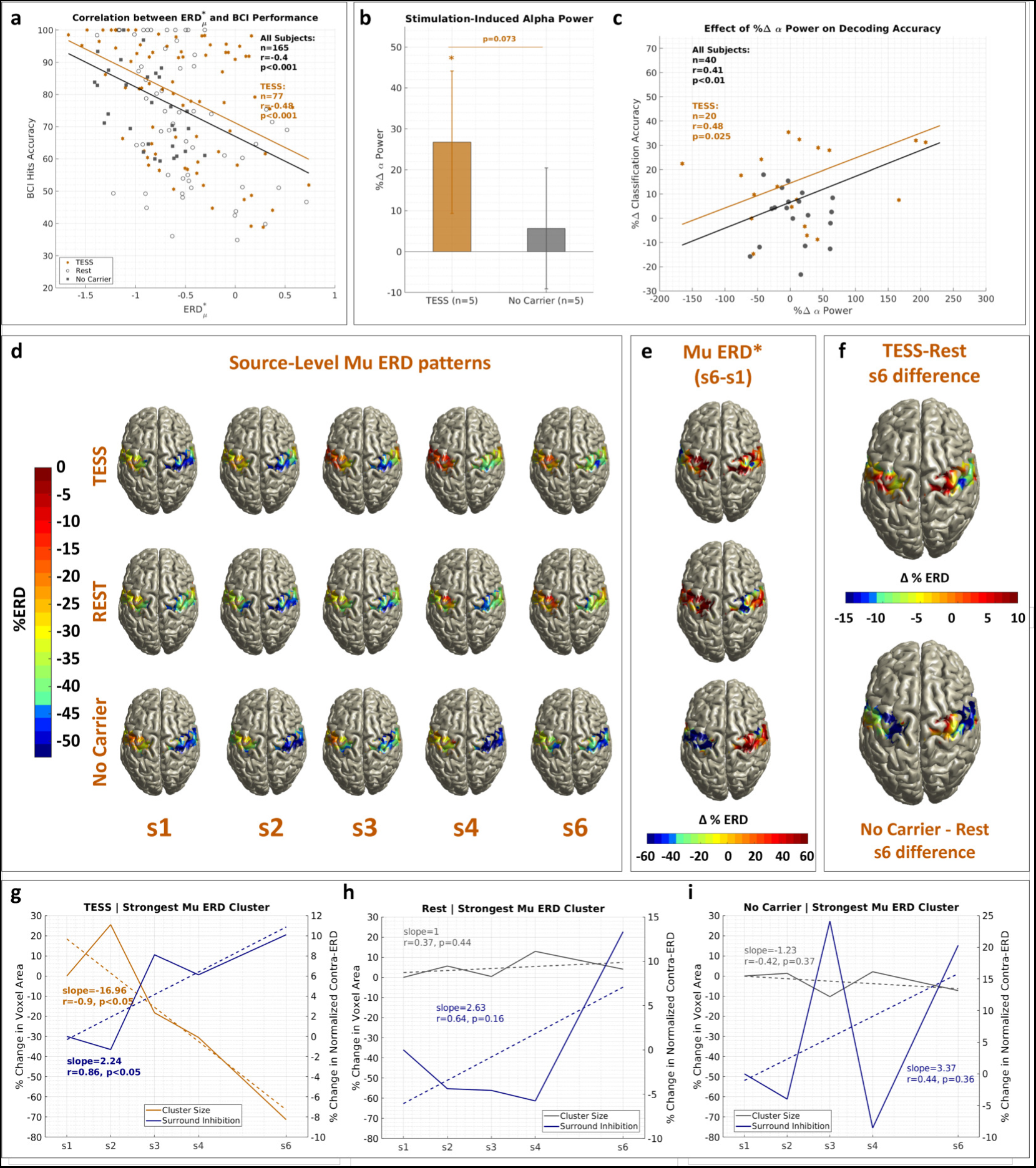
Physiological correlates of enhanced BCI skills. **a**: Correlation between the focality of *µ* ERDs 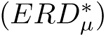 and BCI hits accuracy. The black line corresponds to the line of best fit for all healthy subjects including 14 who had TESS, 10 who had Rest, and 5 who had the no-carrier stimulation. The orange line corresponds to the TESS subjects only. **b**: Percentage change of *α* power post-stimulation for the subset of subjects who had pre- and post-stimulation resting-state EEG recordings. ∗ : *p <* 0.05, absence of asterisks indicates non-significant difference, and error bars represent standard deviations. **c**: Correlation between the percentage change in *α* power and the percentage change in decoding performance post-stimulation for all subjects in black and the TESS group in orange. **d**: Source-level *µ* ERD patterns across the six training sessions for the TESS group in the top row, the Rest group in the middle one, and the No-Carrier group in the bottom row. Contra-lateral ERDs shall appear on the right hemisphere around the hand motor area. **e**: Normalized *µ* ERDs in session 6 within the motor cortex for the TESS, Rest, and No-Carrier groups from top to bottom. **f** : Differences in *µ* ERD patterns at the end of training between the TESS and Rest groups on top and the No-Carrier and Rest groups in the bottom. **g**, **h**, and **i**: Group-level trends in the TESS, Rest, and No-Carrier groups, respectively, for the size of the most significant contra-lateral ERD cluster and its normalized ERD strength relative to the surrounding ERS (Surround Inhibition) in the sensorimotor area. Dotted lines are lines of best fit.

To localize the effects of stimulation in the brain, we reconstructed the source-level ERD patterns for the three groups TESS, Rest, and No-Carrier (Fig. 7d). The TESS group shows an emerging focal ERD over the hand sensorimotor area on the contra-lateral hemisphere. This is particularly evident in Fig. 7e, which shows the normalized ERD values within the entire sensorimotor cortex on the last training session. Fig. 7f shows the difference in raw ERD patterns between the two stimulation groups and the Rest group. When compared to Rest, TESS shows more localized ERDs surrounded by stronger ERS whereas the No-Carrier group exhibits more spread- out ERDs with ipsilateral activation. To quantify the difference in focality, Fig. 7g-i tracks the size of the most significant ERD cluster across sessions and backtracks the normalized strength of ERD (surround inhibition) for the significant cluster on session 6. Only TESS in Fig. 7g shows very strong and significant group-level trends over sessions (Cluster Size: *r* = −0.9*, p <* 0.012; Surround Inhibition: *r* = 0.86*, p <* 0.034). The latter were also strongly and significantly correlated with enhanced classification accuracy (Cluster Size: *r* = −0.88*, p* = 0.022; Surround Inhibition: *r* = 0.90*, p* = 0.015).

## Discussion

Our controlled randomized study is the first to investigate the use of spinal stimulation through cervical TESS in MI-based BCIs. Our results revealed a constructive role for pre-training cortical inhibition in promoting focal and strong SMR modulation as well as BCI skill learning for healthy subjects and for an SCI patient. We validated our results in a longitudinal training framework that tracks skill learning over five days. Supplementary Fig. 6 provides a summary of the grouped results from the different experiments. Starting from comparable baseline to the Rest group, subjects who received TESS achieved significant improvement in BCI performance relative to their baseline after two training sessions only —half of what it took for subjects without stimulation—, and they also performed significantly better at the end of training. More importantly, the trend for the TESS group demonstrated a performance curve that is consistent with the negatively accelerated power law of practice [55] (BCI hits over the 6 sessions: *r* = 0.92*, p <* 0.01*, n* = 6 on log scale). Participants who had TESS also showed consistency and persistence of performance gains, which are essential characteristics for inferring skill learning from performance curves [55, 59]. This can be assessed through retention tests of performance in a setting that is not preceded with further practice but with a break period to allow for the decay of transient temporary influences of stimulation. Session 5 of the protocol in Fig. 1b, which was not preceded by stimulation, serves as an initial retention test for skill learning by evaluating the one-day post-training performance. Supplementary Fig. 6a-c show that TESS subjects maintained significant improvement in all performance metrics on session 5 (*t*_107_ *<* 4.55*, p <* 0.001), and they also maintained the focality of their *ERD_µ_* modulations (Supplementary Fig. 6e, *t*_109_ = −4.03*, p* = 0.001). For the Cross-Over group, a stronger one-week post-training retention test also showed maintained BCI performance improvement and focal modulations (Fig. 4a-c). Incidentally, we did not observe significant short-term effects of stimulation on performance —neither at the beginning of the protocol (session 1 to 2) nor at the end (session 5 to 6). A possible explanation is that TESS is not temporarily modulating neural patterns in a way that is easier to decode, but it is rather activating mechanisms that aid in the longitudinal learning process by which users acquire the MI skill. Drawing analogies to motor learning, which can be defined as a process that uses practice to generate a strong and persistent capability to move skillfully (habit) [55], learning the MI skill can be thought of as a process that increases the capability of vividly retrieving a specific motor program —and its associated kinesthetics— from memory. The latter results in SMR modulations in the sensorimotor area of the involved limb, similar to those observed for actual movements [20]. Our results highlight a role for TESS-induced cortical inhibition in promoting the capability to produce strong and focal volitional SMR modulations, thus accelerating MI skill learning.

Studying the cortical effects of TESS in healthy subjects and SCI patients, Benavides et al. [45] observed an increase in SICI, which is indicative of the activation of GABA inhibitory circuits in the primary motor cortex [60]. The authors further verified that the 5kHz carrier frequency of the stimulation sequence contributed to the cortical inhibitory effects by showing no change in SICI for the no-carrier stimulation [45]. This is also consistent with evidence showing that high frequency (100 Hz) electrical stimulation of skin afferents increases inter-hemispheric inhibition [61], which plays an important role in the generation of proper voluntary movements [62]. For instance, during unilateral hand movements, the active contra-lateral M1 area exerts an inhibitory influence on the ipsi-lateral M1 through pyramidal neurons that projects on GABAergic inhibitory inter-neurons in the latter [62, 63]. Since TESS is believed to activate the sensory afferent fibers in the dorsal horn of the spinal cord [37, 41, 45], it might also have similar effects on the cortical GABAergic inhibitory mechanisms. Previous studies have postulated a role for inhibition during MI [49, 50, 53], either through mechanisms that counterbalance the excitatory output of MI to prevent overt movement [49, 50] or that produce surround inhibition as in overt movements [52, 53, 64], which suppresses the retrieval of conflicting motor traces from memory. In our study, we found evidence of cortical inhibition through increased *α* power in resting-state EEG following TESS (Fig. 7b). This was not the case for the group who had no-carrier stimulation (Fig. 7b), which induced cortical excitation [45]. fMRI evidence supports a negative correlation between *α* power and blood-oxygen-level-dependent activation in the sensorimotor cortex and higher order motor control areas [65]. It is also commonly accepted that the increase in *α* synchronization is associated with inhibition in tasks where learned responses shall be withheld or over brain regions that are not task-relevant [58]. One example is how during MI the surround inhibition phenomenon is observed with a focal ERD in the task-relevant contra-lateral areas and surrounding ERS in the task-irrelevant and ipsilateral regions [52, 53, 64]). Interestingly, our results show a significant correlation between the post-stimulation *α* power and the focality of the *µ* ERD during the subsequent BCI session (*r* = −0.58*, p <* 0.001*, n* = 40) supporting our hypothesis that the TESS-induced inhibition promotes focal SMR modulations. Additionally, despite the limitations of source reconstruction, we observed that the focality of the ERD for the TESS modality was localized over the hand-associated sensorimotor area (Fig. 7d-f). This was not the case for the subjects who received no-carrier stimulation; instead, the latter group showed more spread-out ERD patterns that extended to the ipsilateral region hinting for a causal relationship between inhibitory conditioning and the generation of more focal ERD modulations.

Coupling TESS-induced inhibitory conditioning of the motor cortex with closed-loop feedback training promotes the ability of users to selectively modulate relevant sensorimotor areas; i.e., to vividly retrieve the task-relevant motor traces from memory while suppressing irrelevant information. In [66], authors hinted to a similar potential role for inhibition, induced by mindfulness training, in enhancing performance for MI versus Rest BCIs. Mindfulness helped users increase their *α* power during rest tasks [66]. However, contrary to our findings, it did not result in significant differences in electrophysiological activity during MI. This undermines the generalization of their method to BCIs requiring the distinction between MI patterns of different movements. A potential explanation for how the combination of TESS-induced inhibition and longitudinal MI-BCI training contributes to focal activation revolves around the *α* inhibition-timing hypothesis [58], which postulates that *α* oscillations reflect rhythmic changes in the level of membrane-potential depolarization (excitatory/inhibitory) for large populations of neurons [58, 67, 68]. Following [58], Supplementary Fig. 7 illustrates the mechanism of *α* inhibition-timing for a simplistic example in which a certain population of neurons may fire tonically if its excitation level is high enough to overcome inhibition or it may fire rhythmically —entrained to the *α* rhythm— if either its excitation level is low or the amplitude of the *α* oscillation is large. Following TESS, *α* power increases within the motor cortex causing widespread inhibition and rhythmic activity that affects the entire region including the population of neurons that are relevant to the task. However, the intrinsic excitability of the latter neuronal population will increase with longitudinal MI-BCI training due to activity-dependent changes associated with learning [69]. That is, the MI-relevant populations overcome the effect of cortical inhibition induced by TESS while other regions remain suppressed leading to stronger and more localized surround inhibition.

TESS-induced inhibition also helps constrain the neural dynamics during SMR modulation leading to stronger stability in the BCI decoder feature space. The analysis of the electrode discriminancy score (EDS), which quantifies the contribution of EEG channels to the decoding accuracy, shows very stable patterns for TESS (Supplementary Fig. 8a). Compared to Rest and No-Carrier groups, TESS exhibited more consistent and specific contribution from the physiologically relevant C4 and C3 channels over all of the training sessions. In fact, the average correlation between EDS topoplots on consecutive days, which are susceptible to non-stationarity in neural patterns, is higher in TESS (*r* = 0.92) than Rest (*r* = 0.70). Feature stability translates to more contingent and informative feedback during training, which can consequently accelerate the progression through the learning stages of skill acquisition. This can be particularly relevant to slow learners who face difficulty gaining BCI control [70]. Remarkably, our results from the Cross-Over group of slow BCI learners (Fig. 4) illustrate that TESS enabled them to acquire BCI control after they had failed to do so over the course of 6 training sessions without stimulation. More importantly, this group maintained their BCI performance for one week following training with TESS indicating strong retention of the learned skill. Consistently, Supplementary Fig. 8b shows that the EDS patterns became more focal and remained stable over the C4/C3 channels after the latter group trained with TESS —even for the one-week follow-up session. In fact, the group average contribution of C4/C3 channels for this group was significantly increasing over the training sessions (Cross-Over group: *r* = 0.98, *p <* 0.001*, n* = 6).

Learning and consolidation of SMR modulation is also evident in the case of Patient-I who was stimulated above his lesion site. [45] validated the inhibitory effects of TESS over the C5-C6 segments for SCI patients with similar lesion sites. Consistent with such inhibitory effects, our results showed an increase in resting-state *α* power post-stimulation. The latter was strongly correlated with the increase in online decoding accuracy for TESS sessions of this patient (*r* = 0.96*, p* = 0.021*, n* = 4). Patient-II did not show similar improvements. This was partly expected as stimulation was administered below the lesion site. The reason we still recruited the patient was to explore whether TESS could have an effect at the cortical level assuming residual connections through the lesion site. However, we did not observe an increase in *α* ERS post-stimulation (*t*_3_ = 0.28*, p*−0.813*, n* = 4) nor a significant enhancement of *µ* ERD focality (*session* effect: *F*_5_,_26_ = 1.45*, p* = 0.239, *session*×*modality* interaction effect: *F*_5_,_26_ = 0.99*, p* = 0.445). This was consistent with the poor BCI performance results (Fig. 6d-f).

Despite promising findings, our study also has limitations, such as the method for setting the stimulation amplitude for each subject. The peak-to-peak amplitude of the stimulation current was set to the maximum tolerable value that did not result in muscular contractions. Maximizing the stimulation intensity aimed to obtain the previously reported cortical effects of TESS. This variability across subjects might be a confounding factor. However, we did not observe a stimulation dose effect on BCI performance (Classification Accuracy: *r* = −0.04, *p* = 0.833, *n* = 32; Bar Dynamics: *r* = −0.03, *p* = 0.859; BCI Hits: *r* = 0.05, *p* = 0.776, *n* = 32). Another limitation is the lack of a sham stimulation group. Although the latter would have improved the rigor of our study, previous reports [45] showed no modulatory effects of sham stimulation neither for healthy subjects nor for SCI patients. Incidentally, the No-Carrier group in our study serves as a control for placebo effects —in addition to its main significance in probing the specific role of inhibition by testing the opposite excitatory stimulation condition. Subjects in this group were blinded to the type of stimulation and were given the exact same instructions as in the TESS group. This group actually could not acquire BCI control compared to the Rest group (*session* × *group* interaction effect for BCI hits and bar dynamics: *F*_5_,_324_ *>* 3.13, *p <* 0.01), which rules out a non-modulatory placebo effect for stimulation and supports a causal role for inhibition in driving the learning of SMR modulations and BCI control skills.

Methodologically, only Patient-I and subjects of the Cross-Over group had individual MRIs that were used for source reconstruction while a template brain MRI was used for the remaining participants. Although the group-level analysis for the latter subjects showed ERD sources that are consistent with the expected lateral location of the hand motor area, we acknowledge that the use of template-based reconstruction has its limitations given anatomical and physiological differences across subjects. In future work, we plan to accurately track the focality of ERD sources through multiple fMRI recordings while subjects perform the intended task. Another limitation is the lack of elaborate assessment for the inhibitory effects of TESS that has been extensively studied using TMS-based SICI [45]. However, introducing this assessment in our protocol would have required delaying the BCI-training sessions after TESS conditioning with the risk of compromising the premise for our hypothesis as the potential inhibitory effect could have faded out. Instead, we rely on a neurophysiological correlate of inhibition in terms of increased *α* power during resting-state EEG [52, 53, 57, 58, 65, 67, 68].

We have validated our TESS-BCI approach with a standard task of left versus right hand MI because they exhibit well-defined and well-studied ERD patterns [5, 19, 54] that can be tracked to assess inhibition/focality and to identify evidence of skill learning. Also, MI of hand movements is common in BCI-based motor rehabilitation after stroke [12, 13, 21]. Our TESS approach could be extended to BCIs with higher degrees of freedom and more complex MI commands.

In conclusion, our findings advance the BCI field in general and its clinical applications by highlighting an overlooked potential for inhibitory neuromodulation in conditioning the brain. While our study did not explicitly include assessments of functional gains for individuals with motor disability following the BCI intervention, it provided evidence of enhanced and focal SMR modulations, which are commonly correlated with increased CST excitability [21–24]. It also showed significantly faster and more effective learning of BCI control, which can complement and accelerate previously reported functional benefits with BCI-based rehabilitation interventions [11–16, 21]. Indeed, achieving fast and high BCI control is critical not only for reliable operation of assistive devices, but also for fostering activity-dependent plasticity. Yet, plasticity requires contingent BCI feedback and repetitive activation of the same neural populations, which is at odds with the intrinsic non-stationarity of SMR. Our study shows how the use of inhibitory neuromodulation elicits focal and stronger SMR. This holds significant implications for regenerative medicine that relies on BCI interventions to promote recovery by inducing plasticity in targeted brain regions. A future goal is to assess whether our proposed method can indeed promote functional recovery in different conditions of neuromotor impairment.

## Methods

### Subjects

A total of 25 able-bodied healthy subjects with no history of neurological disorders participated in the study (age 18 to 30 years-old, 48% female). In addition, three SCI patients were recruited (2 males, 1 female). The female SCI participant had to be excluded due to substantial lower back pain arising from her recent injury, which prevented her from completing all sessions. The first male SCI participant (referred to as Patient-I) had his injury at the age of 13, nine years prior to the study. He is clinically diagnosed as tetraplegic with a C5 fracture and a complete lesion, and he has been undergoing consistent motor-rehabilitation therapy. The second SCI participant (referred to as Patient-II) incurred his injury within three months of the study at the age of 26, and he is diagnosed as tetraplegic with a complete spinal cord injury involving a C4 fracture and cervical stenosis. Notably, the latter participant assumed a nearly supine position during the experiment to alleviate back discomfort, and he was receiving stimulation at C5/C6 below the lesion site. All participants, both healthy and with SCI, provided written informed consent to the procedures of the study as approved by the University of Texas at Austin Institutional Review Board (IRB protocol number: 2020-03-0073), and they were compensated for their participation. The study protocol is published on ClinicalTrials.gov (NCT05183152) and the CONSORT enrollment flow-diagram is provided in Supplementary Fig. 9. **Study Design.** *Main Group.* In the main part of the study, a cohort of 20 individuals without prior experience in utilizing MI-based BCIs completed the longitudinal BCI training protocol depicted in Fig. 1b. These participants were randomly allocated to either the experimental group, which received pre-training TESS stimulation lasting 20 minutes (referred to as TESS group), or the control group (referred to as Rest group) that engaged in a 20-minute resting period before training. The initial day of the protocol started with an offline BCI session wherein participants perform cued MI for movements of their left or right hand. MI instructions highlighted the importance of performing kinesthetic —not visual— MI by emphasizing the mental rehearsal of the sensation and muscular tension association with the execution of the movement. A Riemannian-geometry-based BCI decoder was trained on the collected offline data and was used to provide real-time feedback during subsequent online sessions. On the second day, participants started with an online BCI session, which serves as their performance baseline. This was followed by 20 minutes of rest or stimulation, depending on their group assignment, and then a post-conditioning online BCI session. After this second day, the BCI decoder was re-calibrated to incorporate data from the post-conditioning online session, and then it was fixed for the remainder of the study. Days 3 and 4 included 20 minutes of conditioning followed by an online BCI session. Finally on Day 5, participants started with an online BCI session, which tests the one-day-after effect without further conditioning, then they completed one last 20 minutes of conditioning followed by the final online BCI session. In addition to healthy participants, patients with SCI completed the protocol of Fig. 1b once with the TESS condition and once with the Rest condition, with a one week separation period. Following random order selection, Patient-I started with TESS then Rest while Patient-II followed the opposite order. The patients also completed 2 minutes of resting state EEG recordings before an after each stimulation session.

*Cross-Over Group.* This subgroup includes the participants from the Rest condition who were unable to attain BCI control by the end of the study. A total of four subjects exhibited chance-level performance consistently across all of the training sessions. These participants were subsequently re-enlisted to undergo the TESS condition, adhering to the same protocol of Fig. 1b with the addition of 2-minutes of resting state EEG before and after each stimulation session. The washout period for this group was six months to ensure no carry-over effect from the initial training. In order to comprehensively evaluate the impact of TESS on enhancing their learning capabilities, we performed an assessment of their capacity to sustain the attained level of performance during a session occurring one week later. Importantly, this evaluation was conducted without any supplementary TESS stimulation, thus further illuminating the enduring effects of TESS-induced skill enhancement.

*No-Carrier Group*. This subgroup includes an additional five participants who underwent the same training protocol with a distinct stimulation condition, closely resembling TESS but lacking the 5kHz carrier frequency (Fig. 1a). Specifically, it consists of a single 200us biphasic pulse within one period. This specific stimulation sequence is believed to elicit excitatory effects at the cortical level, contrary to the inhibitory effects brought about by the conventional TESS sequence [45]. This group also completed 2 minutes of pre- and poststimulation resting state EEG recordings. The inclusion of this group serves a twofold purpose: firstly, it effectively controls for potential placebo effects, ensuring the discernment of genuine outcomes; and secondly, it substantiates the premise that TESS inhibitory facet is the pivotal driving force behind the observed enhancements in SMR modulation.

### BCI Training

*Bar Task.* The MI-based BCI training employs a conventional bar feedback task [5]. In offline sessions, the bar moves consistently to either the right or left direction over the duration of the task execution depending on the class of the trial (i.e., right hand or left hand MI). In online sessions, the bar provides real-time feedback on MI performance. It moves according to an accumulated evidence of posterior probabilities generated by the binary-class BCI decoder, which classifies EEG signals into one of the two classes. The BCI decoder operates on samples of a 1-second sliding window with a step size of 62.5ms, and it accumulates evidence for MI classes according to the following equation:

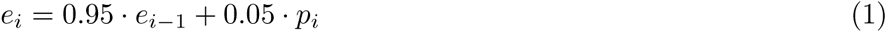

where *e_i_* is the accumulated evidence for a given class and *p_i_* is the posterior probability of that class for the *i^th^* sample. *e_i_* is reset to 0.5 at the beginning of the task execution period of every trial, and a trial ends when *e_i_* crosses a predefined command delivery threshold for either class. The outcome scenarios of a trial are: “trial hit,” where a correct command is delivered upon *e_i_* crossing the threshold on the correct side aligned with the trial’s class; “trial miss,” where an incorrect command is delivered if *e_i_* crosses the threshold on the wrong side; and “timeout,” when neither threshold is crossed within a 7-second period. For each run, threshold values are adjusted to provide a suitable challenge level, maintaining participant engagement while minimizing the likelihood of erroneous commands and optimizing the probability of accurate commands.

*Session Structure.* A session consists of 3-4 runs, each with 20 trials that are randomly split into right or left hand MI (see Fig. 1d). A trial starts with a rest period of 2 s with a random jitter of 1 s, followed by a fixation period for 2 s, a left or right arrow cue indicating the MI task for 1.5 s, a task execution period of 5 s in offline runs and a timeout duration of 7 s in online runs, and a trial result feedback of 2 s.

### EEG/EOG

EEG was recorded at 512 Hz using an ANTNeuro EEGO amplifier with a waveguard EEG cap (ANT Neuro, Netherlands). The cap uses Ag/AgCl coated soft polymer electrodes positioned over 32 standard scalp locations according to the international 10-10 system. For patient recordings and for the two additional subgroups (Cross-Over and No-Carrier), a higher density 64-channel waveguard EEG cap was used. In both cases, the ground and reference electrodes were placed at AFz and CPz, and the same subset of channels was used for online classification. Electrode impedances were kept below 20 kΩ. Synchronized electrooculography (EOG) signals were collected via a bipolar box connected to the amplifier, from three channels placed on the eye canthi and the forehead with the ground and reference on the mastoids. EOG was used to reject samples with eye blinks or left/right eye movements. All processing and analysis of EEG and EOG signals were conducted using MATLAB and the Fieldtrip toolbox [71].

### BCI Decoder

*Feature Extraction.* The real-time EEG classification (right hand versus left hand MI) is performed according to the Riemannian geometry framework, which has been attracting growing interest in non-invasive BCI research [72, 73]. Riemannian classifiers use covariance matrices of mulit-channel EEG as features to characterize spatio-spectral neural patterns. In our processing pipeline, trace-normalized covariance matrices in equation (2) are estimated for a subset of 22 EEG channels (i.e., F7, F3, Fz, F4, F8, FC5, FC1, FC2, FC6, C3, Cz, C4, CP5, CP1, CP2, CP6, P7, P3, Pz, P4, P8 and POz) using the shrinkage-based covariance estimator method of Ledoit and wolfe [74] after bandpass filtering EEG to [8, 30] Hz using a 2nd order causal Butterworth filter.

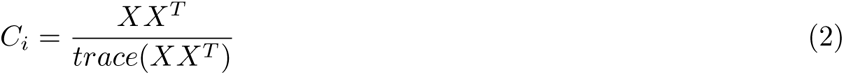

where *C* is the sample-wise normalized covariance, *X* ∈ ℝ*^Nc×Nt^* denotes a 1-second EEG sample, *N_c_* is the number of channels, and *N_t_* is the number of time points. Since covariance matrices are symmetric positive definite (SPD), they lie on a Riemannian manifold, whose geometrical properties allow defining the geodesic distance *δ_r_*(*C*_1_*, C*_2_) between two SPD matrices *C*_1_ and *C*_2_ according to the following equation [72]:

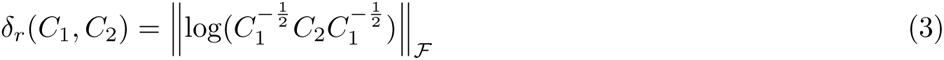

*MDM Classifier.* For classification purposes, class prototypes (for left hand versus right hand MI) are constructed as the centers of mass C̄ (or karcher means) for their respective features *C_i_* on the Riemannian manifold by solving the optimization problem in equation (4) over the space of *N_c_*-dimensional SPD matrices *P_n_* using the method outlined in [72].

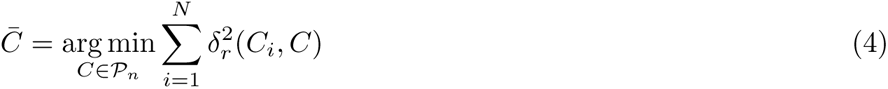

Using the estimated class prototypes and the geodesic distance metric of equation (3), a simple Minimum Distance to Mean (MDM) classifier can be used to classify an EEG sample based on the shortest geodesic distance between its covariance matrix and the class prototypes. In an online setting, the classifiers output is blocked whenever an EOG artifact is detected.

### TESS

We used the same stimulation sequences and settings as in [45] to ensure we elicit similar modulatory effects. TESS was delivered for 20 minutes over the neck at the cervical level between the C5-C6 spinal segment and the iliac crests of the hip bone as depicted in Fig. 1a. A 3.2-cm-diameter hydrogel adhesive electrode was used on the neck and two 7.5x12 cm rectangular electrodes (ValuTrode Cloth, Axelgaard Manufacturing) were located bilaterally over the iliac crests. We used a Hasomed Rehamove3 stimulator to generate the stimulation sequence of 200 us biphasic pulses at a frequency of 30 Hz with a carrier frequency of 5 kHz as depicted in Fig. 1. The peak-to-peak amplitude of the stimulation current was set to the maximum tolerable value that did not result in any muscular contraction (14 healthy: 41.0 ± 9.5 mA, Patient-I: 44 mA, Patient-II: 40 mA). For the No-Carrier group, TESS without 5 kHz carrier frequency was delivered for 20 minutes with single biphasic pulses (200 us duration) at a 30 Hz frequency. This paradigm was tolerated by all subjects, allowing us to match similar stimulation intensity as used in the original TESS protocol (5 healthy: 55.8 ± 12.8 mA). Since changes in pulse duration can affect the relative recruitment of sensory axons, we maintained the waveform pulse duration consistent across paradigms similar to [45].

### Statistical Analysis

We used linear mixed effect models (LMEs) to perform statistical testing and regression analysis. Specifically, LME-ANOVA was used to test for the main and interactions effects with the between-subjects fixed effect as the group condition (TESS, Rest, TESS w/o 5 kHz carrier), the within-subject fixed effect as sessions (sessions 1 to 6 plus follow-up sessions), and the random effect of subjects (to account for paired comparisons). Appropriate LME models were used for post-hoc paired and independent tests, and p-values were corrected with the Holm-Bonferroni method for multiple comparisons. We verified the normality of the residuals with the Lilliefors test, and whenever it is violated, we used Mann-Whitney U test for paired samples (U statistic reported), Wilcoxon signed-rank test for independent samples (Z statistic reported), and Spearman’s rank correlation coefficient for correlation analysis. For studying the learning curve from the BCI performance over sessions, we identified the inflection point as the session that minimizes the residual error of horizontal lines of best fit from the pre and post inflection segments, and we parameterized a piece-wise linear spline function around the inflection to characterize the period of significant steep ascent and the one with plateau. Regression lines for performance metrics over sessions were fitted using the medians across subjects in order to track progressive group-level improvements even when the values saturated at 100% accuracy. We used cluster permutation testing —implemented in FieldTrip— to correct for multiple comparisons in spatio-temporal (time-frequency plots) or spatio-spectral (topoplot or source space) analysis. Neighboring electrodes, voxels, frequency bins, or time instances that show sample-level significant effects with a t-test are grouped into clusters. The sum of the t-statistic is computed within each cluster to form a cluster-level statistic. The test statistic is defined as the maximum of the cluster-level statistic values over all detected clusters. The reference distribution for the test statistic is computed through the Monte Carlo method by randomly permuting the observations across subjects (between-subjects design) or across the spatio-temporal or spatio-spectral dimensions. The significance probability (p-value) is then computed for the observed test statistic based on the approximated reference distribution.

### Outcome Measures

*BCI Performance*. The BCI performance is assessed by the following metrics:

- Sample-wise Classification Accuracy: It represents the online decoder accuracy for the 1*s* overlapping BCI samples, and it is computed as follows:

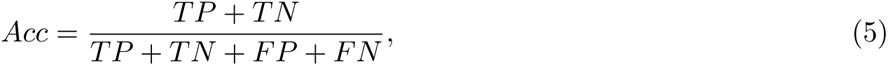

where *TP*, *TN*, *FP* and *FN* correspond to true positives, true negatives, false positives and false negatives, respectively.
- Bar Dynamics: It is a measure of BCI control, and it represents the percentage of time during the MI task execution period when the user could keep the feedback bar on the correct side of the interface. Bar dynamics track the accumulated evidence of the BCI decoder.
- Command Delivery/BCI Hits: It measures the accuracy with which the feedback bar reaches the command delivery thresholds. It is the ratio of correctly delivered commands to the total commands delivered. This metric shall be analyzed along with the timeout ratio, which accounts for the percentage of trials in which no command was delivered.

*Electrode Discriminancy Score (EDS).* We use the recently proposed EDS for the Riemanian geometry classification framework [32] to quantify the contribution of each EEG channel to the BCI decoding accuracy. The metric assesses channel contributions through a backward elimination approach, which quantifies the loss in posterior prediction accuracy following the removal of a given channel from the model.

*Physiological*. We track the evolution of ERDs over the course of training to test how different conditions affect the evolution of ERD patterns. Particularly, we assess the strength and focality of ERDs by identifying the EEG channel that exhibited the strongest ERD (*ERD*^∗^) on the last training session, and we backtrack its values across sessions. To test for the focality of the ERD at that channel, we normalize its value to the ERDs within the motor area *ERD_motor_* as in equation (6) or just within the contra-lateral motor area.

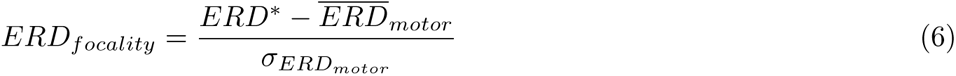

### ERD/ERS

Trial periods with EOG artifacts were rejected from analysis. To compute ERD/ERS patterns, EEG signals were bandpass filtered to the frequency range of interest (*µ*, ([8, 13] Hz), or *β*, ([18, 30] Hz)) using a non-causal 4*^th^* order Butterworth filter, spatially filtered using a Laplacian plus montage, squared to compute the instantaneous power, and then averaged over the analyzed period. The average power in the MI execution period was normalized to a pre-task [-0.8, 0] s interval —avoiding any residual visually evoked potentials from cue presentation.

### Resting State EEG

We computed the power of the *α* band oscillations ([8, 13] Hz) as a correlate of cortical inhibition, and we wanted to quantify how TESS might modulate resting state *α* oscillations. We used irregular resampling auto-spectral analysis (IRASA) [75], a method used to distinguish between rhythmic activity and concurrent power-spectral 1/f modulations. This technique involves modifying the time-domain data through a series of non-integer re-sampling factors before conducting Fourier-based spectral decomposition. As a result, the powerspectrum’s rhythmic components are redistributed, while the non-rhythmic 1/f distribution remains unchanged. By calculating the median of the resulting auto-spectral distributions, the power-spectral 1/f component can be isolated and subtracted from the original power-spectrum allowing us to estimate the power-spectral contribution of rhythmic oscillations in the recorded signal.

### MRI

MRI images were obtained for Patient-I and for the participants from the Cross-Over group on a 3T whole body scanner (Magnetom Vida, Siemens, Germany) using a standard head coil. Subjects’ heads were immobilized with a vacuum pad to reduce movement artifacts. For each subject, an anatomical volume data set was acquired using a T1-weighted 3D-MPRAGE sequence (TR 2400 ms, TE 2.18 ms, flip angle 8°, voxel size 0.8x0.80.8 *mm*^3^s).

### Source-Level Analysis

We followed the analysis pipeline detailed in [76] using Fieldtrip. We used frequency-domain beamformers based on the dynamical imaging of coherent sources (DICS) method, which makes use of the cross-spectral density matrix of the data and a forward model of the brain to estimate the spatial filters necessary for source reconstruction. For Patient-I and the participants in the Cross-Over group, we obtained T1-weighted MRI images to construct volume conduction models and build subject-specific forward models. For other participants, we used a realistically shaped three-layer boundary-element template volume conduction model on a 3D grid of dipole locations with equidistant spacing of 8mm, and EEG channel locations were co-registered to the used conduction models. The choice of the frequency of interest for the DICS method was guided by time-frequency analysis within the frequency bands of interest.

## Author Contributions

H.A. and J.d.R.M. conceived and designed the experimental protocols. H.A. implemented the experimental protocols. H.A., D.L., and J.M. were responsible for data acquisition. H.A., D.L., and S.K. performed the analysis. H.A., D.L., J.M., S.K., F.S.R. and J.d.R.M. interpreted results. All authors prepared the draft manuscript. All authors reviewed the results and approved the final version of the manuscript.

## Data Availability

All data produced in the present study are available upon reasonable request to the authors.

## Acknowledgments

This work was partially supported by the Coleman Fung Foundation.

## Competing Interests

Authors declare no competing interests.

## Notes

### Competing Interest Statement

The authors have declared no competing interest.

### Clinical Trial

NCT05183152

### Clinical Protocols

https://clinicaltrials.gov/study/NCT05183152

### Author Declarations

IRB of The University of Texas at Austin gave approval for this work (IRB protocol number: 2020-03-0073), and the study protocol is published on ClinicalTrials.gov (NCT05183152)

