## Supplementary Material for "Transcutaneous Electrical Spinal Cord Stimulation Promotes Focal Sensorimotor Activation that Accelerates Brain-Computer Interface Skill Learning"

**June 10, 2024**

#### Supplementary Material

##### Slow BCI Learners

###### Performance in Rest protocol

Slow BCI learners were identified in the Rest group as the set of subjects who had around 50% bar dynamics accuracy —indicating chance level control of the BCI. Four subjects were identified, and their performance is depicted in Supplementary Figure 1 over the six training sessions of the protocol. This group had no significant change in their bar dynamics accuracy at the end of training (session 1 $\mu \pm \sigma$: $47.27 \pm 9.33\%$, session 6 $\mu \pm \sigma$: $47.40 \pm 8.77\%, t_{27} = 0.038, p = 0.970$).

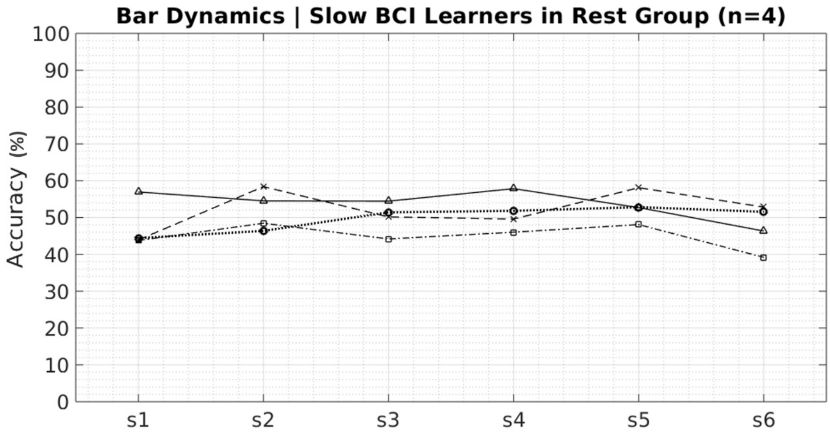


Supplementary Figure 1: **Slow BCI learners.** The variation of bar dynamics accuracy over the six training sessions for the set of slow BCI learners within the Rest group who could not reliably control the BCI. Bar dynamics of 50% indicates chance level.

###### Analysis of $\beta$ ERD

Supplementary Figure. 2 compares $\beta$ ERDs in the group of slow learners for the Rest and TESS protocols —after a six-month washout period. The figure backtracks the $ERD_\beta$ for the channel with the strongest desynchronization on the last training session. While none of the modalities (Rest or TESS) show significant differences in the magnitude of $ERD_\beta$ at the end of training, the results show a significant main effect of the $session$ on the $ERD_\beta$ focality ($session$ ($F_{5,168} = 2.49, p = 0.033$, Supplementary Figure. 2c) with both modalities showing significant trends of enhanced focality (Supplementary Figure. 2c, Rest: $slope = -0.12, r = -0.3, p < 0.01$, TESS: $slope = -0.08, r = -0.26, p < 0.05$).

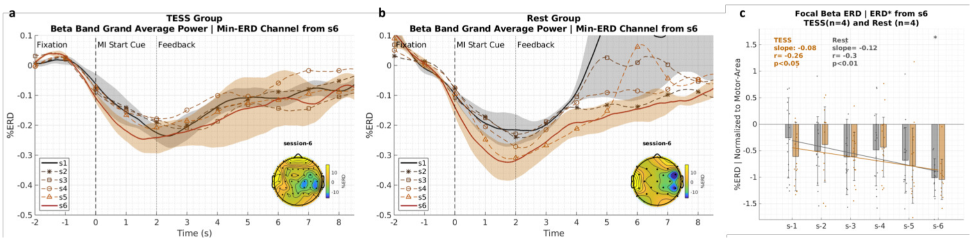


Supplementary Figure 2: **Beta ERD analysis for the group of slow BCI learners.** **a**, **b**: Grand average $\beta$ ERD patterns for the channel that showed the highest ERD strength at the end of training similar to Fig. **??**a,b for the TESS (orange) and Rest (gray) modalities, respectively. **c**: Focality of $\beta$ ERD patterns ($ERD^*$) for the TESS (orange) and Rest (gray) groups similar to Fig. **??**g. $*p < 0.05, **p < 0.01, ***p < 0.001$, absence of asterisks indicates non-significant difference. All p-values are Bonferroni-Holm corrected for multiple comparisons.

### Supplementary Material: Transcutaneous Electrical Spinal Cord Stimulation Promotes Focal Sensorimotor Activation that Accelerates Brain-Computer Interface Skill Learning

✉

**June 10, 2024**

#### Source-level $\mu$ ERD analysis

Supplementary Figure 3 details the source-level analysis of the $ERD_\mu$ for the group of slow learners when they completed both Rest and TESS protocols. Only with TESS a stronger contra-lateral $ERD_\mu$ emerges on the right hemisphere over the course of training. On the last sixth session, the difference between source-level patterns show a stronger contra-lateral desynchronization —accompanied with larger ipsi-lateral synchronization— for TESS relative to Rest. This is consistent with our hypothesis that TESS promotes stronger and more focal SMR modulations through inhibitory conditioning of the brain prior to selective cortical modulations during BCI training.

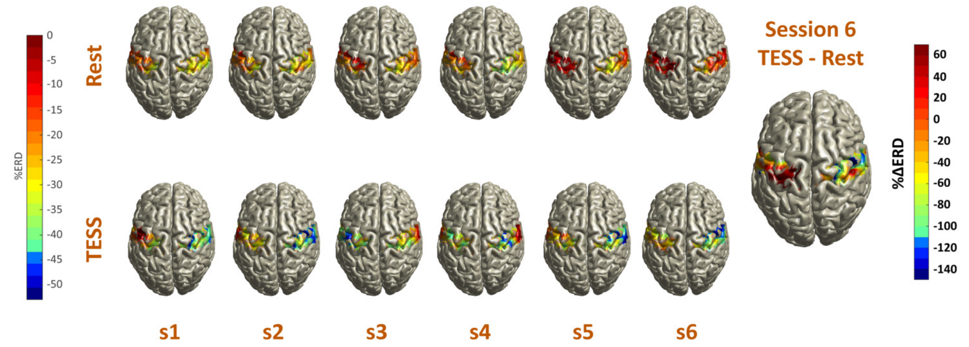


Supplementary Figure 3: **Session-wise source analysis for $\mu$ ERD in the slow learners group.** Top row shows $\mu$ ERD patterns over the sessions of the no-stimulation modality while the bottom row shows the results with TESS. The right-most plot shows the difference in $\mu$ ERD patterns between the two modalities on the last session of training.

**Supplementary Material: Transcutaneous Electrical Spinal Cord Stimulation Promotes Focal Sensorimotor Activation that Accelerates Brain-Computer Interface Skill Learning**

✉

**June 10, 2024**

#### Patient-I ERD Analysis

Patient-I completed both TESS and Rest protocols starting with TESS, with one week of break between the two modalities. Supplementary Figure 4a,b show the change in focality of $\mu$ and $\beta$ ERDs, respectively, over the training sessions. LME-ANOVA reveals a significant interaction effect of $session \times group$ in both bands ($F_{5,31} \geq 7.78, p < 0.001$). Starting with TESS, the figure shows a significant trend of enhanced focality from session 3 ($\mu$: $slope = -0.77, r = -0.78, p < 0.001$, $\beta$: $slope = -0.40, r = -0.78, p < 0.01$), when the decoder was recalibrated with emerging patterns from the first post-stimulation online session (session 2). After completing the TESS protocol, the strength of ERD focality was maintained one week later during the Rest protocol with non-significant changes over subsequent sessions.

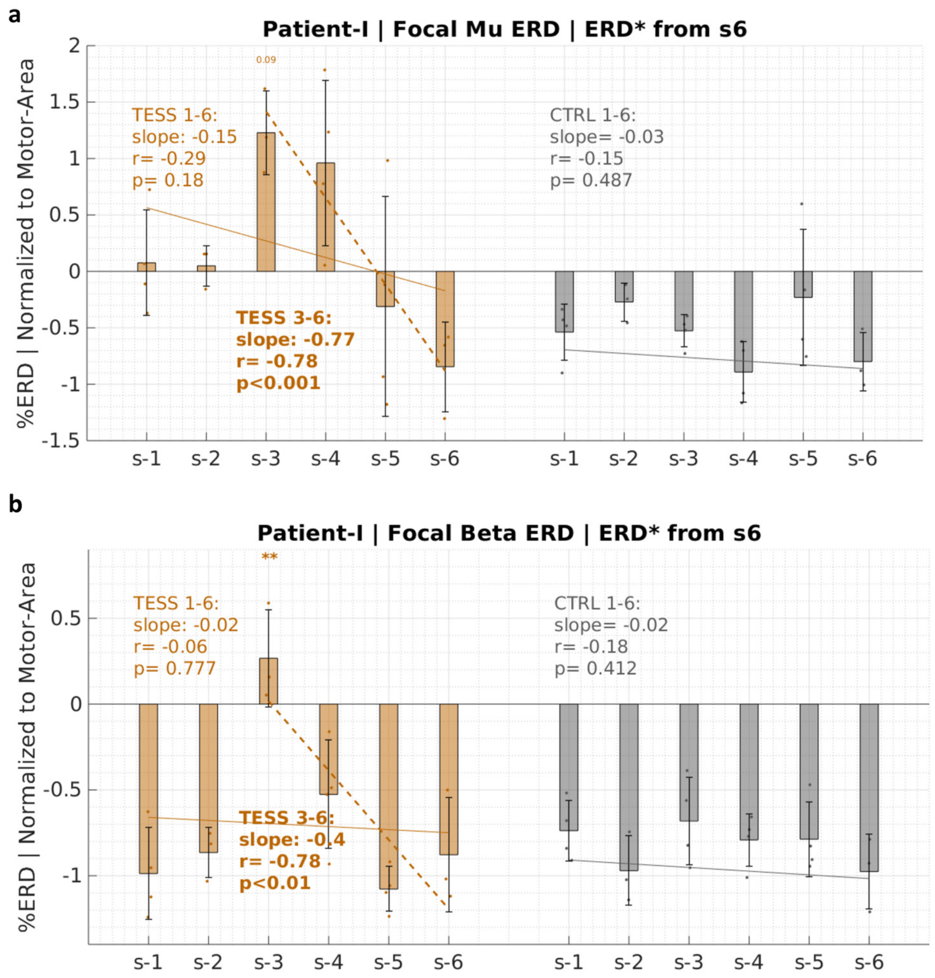


Supplementary Figure 4: **Physiological evidence for SCI Patient-I. a**, **b**: Focality of ERD patterns in the $\mu$ and $\beta$ bands, respectively. The plots backtracks the ERD strength of the channel that exhibited the greatest desynchornization at the end of training ($min - ERD$) after normalizing it to the ERD values within the motor area (i.e., the FC, C, and CP electrodes). Trend lines are regression fits of session means. Within-group significant difference relative to baseline is indicated above the violins with orange asterisks for TESS and gray asterisks for Rest. Statistical testing is performed using linear mixed effect models as detailed in Methods. $*p < 0.05, **p < 0.01, ***p < 0.001$, absence of asterisks indicates non-significant difference. Error bars represent standard deviations.

### Supplementary Material: Transcutaneous Electrical Spinal Cord Stimulation Promotes Focal Sensorimotor Activation that Accelerates Brain-Computer Interface Skill Learning

**June 10, 2024**

#### Correlation between $\alpha$ Power and $ERD_{\mu}^{*}$

Supplementary Fig. **??** shows that the strength of $\alpha$ power post-stimulation explains the increased focality of $\mu$ ERD in the subsequent BCI training session (Spearman correlation, $r = -0.58, p < 0.001, n = 40$). The figure includes results for the Cross-Over group ($N = 4$), Patient-I, and the No-Carrier group ($N = 5$).

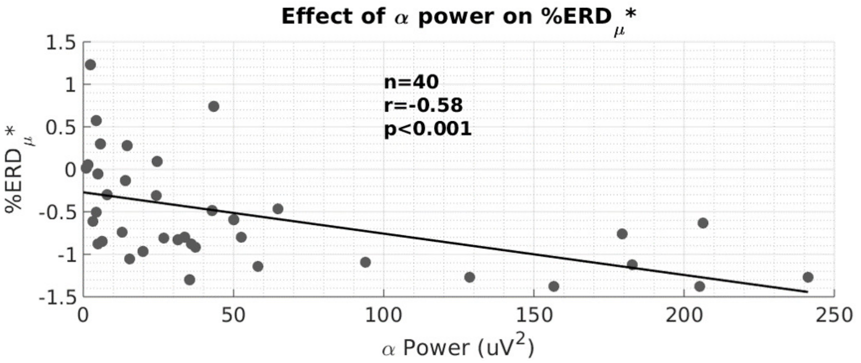


Supplementary Figure 5: **Effect of Post-Stimulation $\alpha$ Power on $\mu$ ERD**

### Supplementary Material: Transcutaneous Electrical Spinal Cord Stimulation Promotes Focal Sensorimotor Activation that Accelerates Brain-Computer Interface Skill Learning

**June 10, 2024**

#### Summary of TESS versus Rest Results for Healthy Subjects

##### BCI performance metrics

With the re-enrollment of the four slow BCI learners from the Rest group to the TESS group, a total of 14 healthy subjects completed the protocol with TESS and 10 healthy subjects with Rest. A summary of BCI performance metrics and ERD focality results for all these healthy subjects is presented in Supplementary Figure 6. LME-ANOVA reveals significant $session \times group$ interaction effects for all three BCI performance metrics (classification accuracy: $F_{5,534} = 2.46, p = 0.032$; bar dynamics: $F_{5,534} = 2.36, p = 0.040$; BCI hits: $F_{5,514} \geq 3.50, p < 0.01$) as well as significant main effects for $session$ (classification accuracy: $F_{5,534} = 5.33, p =< 0.001$; bar dynamics: $F_{5,534} = 3.84, p =< 0.01$; BCI hits: $F_{5,513} \geq 3.83, p < 0.01$). Post-hoc paired tests show significant improvement for the TESS group starting on session 3 for all three metrics ($t_{110} > 4.19, p_{corrected} < 0.001$), where cross-group significant differences also emerged with TESS outperforming Rest ($t_{94} > 2.68, p < 0.02$). The latter remained significant for BCI hits at the end of training ($Z = 2.48, p_{corrected} = 0.026$). Remarkably, the trend in BCI hits for the TESS group demonstrated a performance curve that is consistent with the negatively accelerated power law of practice [1, 2] (BCI hits over the 6 sessions: $r = 0.82, p = 0.026, n = 6$ on log scale). Timeouts were generally less in TESS relative to Rest as depicted in Supplementary Figure 6d.

##### ERD analysis

Supplementary Figure 6d,e compare the change in focality of the $\mu$ and $\beta$ ERDs, respectively, over the training sessions between the TESS and Rest modalities. LME-ANOVA reveals significant $session \times group$ interaction effect for $ERD^*_\mu$ ($F_{5,533} = 2.34, p = 0.040$) with post-hoc tests showing significant cross-group differences at the end of training ($Z = 2.84, p = 0.018$). Only TESS subjects exhibited a significant trend of enhanced $ERD^*_\mu$ focality with training (TESS: $slope = -0.10, r = -0.29, p < 0.001$). In the $\beta$ band, both modalities showed significant trends of enhanced $ERD^*_\beta$ focality with training (TESS: $slope = -0.09, r = -0.27, p < 0.001$, Rest: $slope = -0.08, r = -0.21, p < 0.01$) leading to a significantly stronger $ERD^*_\mu$ at the end of training relative to baseline (Rest: $t_{71} = 4.77, p_{corrected} < 0.001$, TESS: $Z = 5.30, p_{corrected} < 0.001$).

### Supplementary Material: Transcutaneous Electrical Spinal Cord Stimulation Promotes Focal Sensorimotor Activation that Accelerates Brain-Computer Interface Skill Learning

✉

**June 10, 2024**

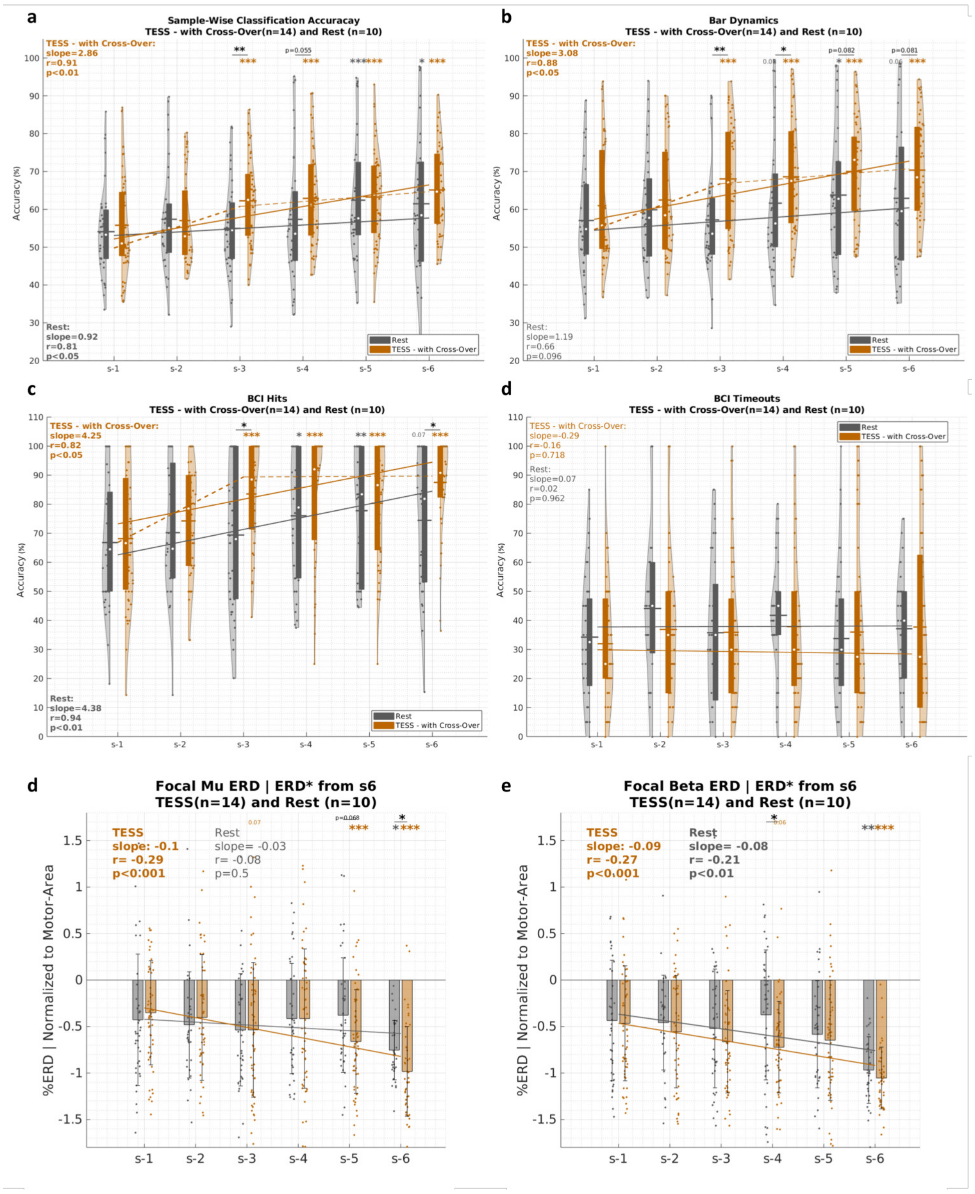


Supplementary Figure 6: **Summary of results for all TESS and Rest subjects. a**, **b**, **c**, and **d**: Classification accuracy, bar dynamics, BCI hits, and timeouts over the six training sessions of the protocol in Fig. **??**b for TESS (orange) and Rest (gray) groups. Boxes within violins correspond to the inter-quartile range, white circles correspond to medians, and horizontal lines correspond to means. Session 1 performance serves as a baseline for within-group comparisons. The solid regression lines correspond to the trends of the medians over sessions. The dashed lines correspond to piece-wise spline regression of medians. Significant differences across groups are indicated by underlined asterisk, and within-group significant difference relative to baseline is indicated above the violins with orange asterisks for TESS and gray asterisks for Rest. **e**, **f**: Focality of $\mu$ and $\beta$ ERD patterns ($ERD^*$), respectively, for the TESS (orange) and Rest (gray) groups similar to Fig. **??**g,h. The solid regression lines correspond to the trends of the means over sessions. Statistical testing is performed using linear mixed effect models as detailed in Methods. $* : p < 0.05, ** : p < 0.01, *** : p < 0.001$, absence of asterisks indicates non-significant difference. All p-values are Bonferroni-Holm corrected for multiple comparisons. Error bars represent standard deviations.

### Supplementary Material: Transcutaneous Electrical Spinal Cord Stimulation Promotes Focal Sensorimotor Activation that Accelerates Brain-Computer Interface Skill Learning

**June 10, 2024**

#### Alpha Inhibition-Timing Hypothesis

One potential explanation for how the combined effects of Transcranial Electrical Stimulation (TESS)-induced inhibition and longitudinal Motor Imagery-based Brain-Computer Interface (MI-BCI) training contribute to focused activation involves the alpha inhibition-timing hypothesis [3]. This hypothesis suggests that alpha oscillations reflect rhythmic alterations in the depolarization levels (excitatory/inhibitory) across large groups of neurons [3–5]. Supplementary Figure 7 —based on [3]—, illustrates how alpha inhibition-timing operates. A specific group of neurons may consistently fire if their excitation level surpasses inhibition, or they may fire rhythmically —synchronized with the alpha rhythm— when either their excitation level is low or the amplitude of the alpha oscillation is significant. After TESS application, there is an increase in alpha power within the motor cortex, leading to widespread inhibition and rhythmic activity affecting the entire region, including the neuronal population relevant to the task. However, with longitudinal MI-BCI training, the intrinsic excitability of these relevant neuronal populations increases due to activity-dependent changes associated with learning [6]. Consequently, these populations override the cortical inhibition induced by TESS, while other regions remain suppressed, resulting in stronger and more localized surround inhibition.

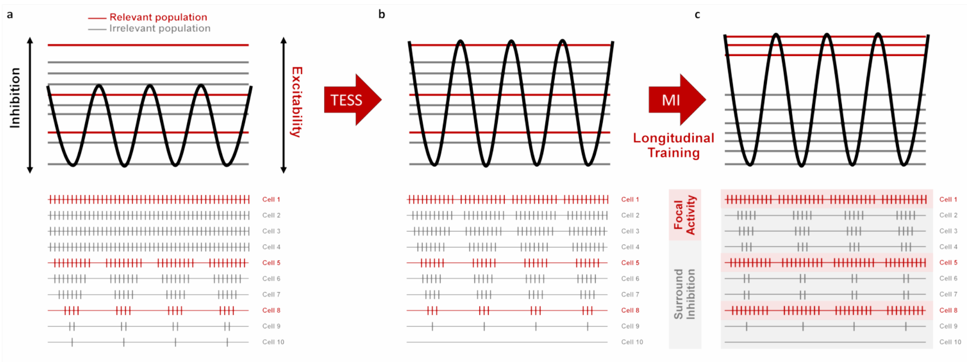


Supplementary Figure 7: **TESS and the $\alpha$ inhibition-timing hypothesis — modified from [3].** An illustrative example for the inhibitory mechanism of the $\alpha$ rhythm. **a**: A certain population of neurons may fire tonically if its excitation level is high enough to overcome inhibition or it may fire rhythmically —entrained to the $\alpha$ rhythm— if either its excitation level is low or the amplitude of the $\alpha$ oscillation is large. **b**: Following TESS, $\alpha$ power increases leading to stronger and widespread inhibition and rhythmic activity within the motor cortex —including the task-relevant population of neurons. **c**: Longitudinal MI training following each session of TESS increases the intrinsic excitability of the task-relevant neuronal population allowing it to overcome inhibition while other regions are suppressed.

#### Supplementary Material: Transcutaneous Electrical Spinal Cord Stimulation Promotes Focal Sensorimotor Activation that Accelerates Brain-Computer Interface Skill Learning

✉

**June 10, 2024**

##### Feature Space Analysis: Electrode Discriminancy Scores

Supplementary Figure 8a,b,c show the topoplot visualizations of z-transformed electrode discriminancy score (EDS) values for the group of healthy subjects (14 for TESS, 10 for Rest, and 5 for No-Carrier), the Cross-Over group of slow BCI learners, and Patient-I, respectively. EDS values show the contribution of each of the used EEG channels to the overall classification accuracy of the BCI decoder. The results in Supplementary Figure 8a show that TESS exhibited more consistent and specific contribution from the physiologically relevant C4 and C3 channels over all of the training sessions. This is reflected in higher correlations for EDS topoplots across sessions in TESS relative to Rest especially for sessions 2 to 5, which occurred on separate days and were more likely to experience higher non-stationarity in neural patterns (TESS: $r_{avg} = 0.92$, Rest: $r_{avg} = 0.70$). In Supplementary Figure 8b, the Cross-Over group exhibit similar characteristics where the slow BCI learners had consistent contributions from the C4 channel upon training with TESS but not Rest. A similar EDS topoplot is also observed in the one-week follow-up session for the TESS modality of the latter group. For the case of Patient-I, the contribution from the contralateral hemisphere emerged on session 2 after a single session of TESS and remained till the end of training. One week later, the EDS topoplots showed consistent contributions to those of the last session of TESS when Patient-I continued the study without further stimulation. Overall, these results support a role for TESS in promoting greater stability in the feature space, which translates to more contingent and informative feedback during training; therefore, accelerating the progression through the learning stages of skill acquisition.

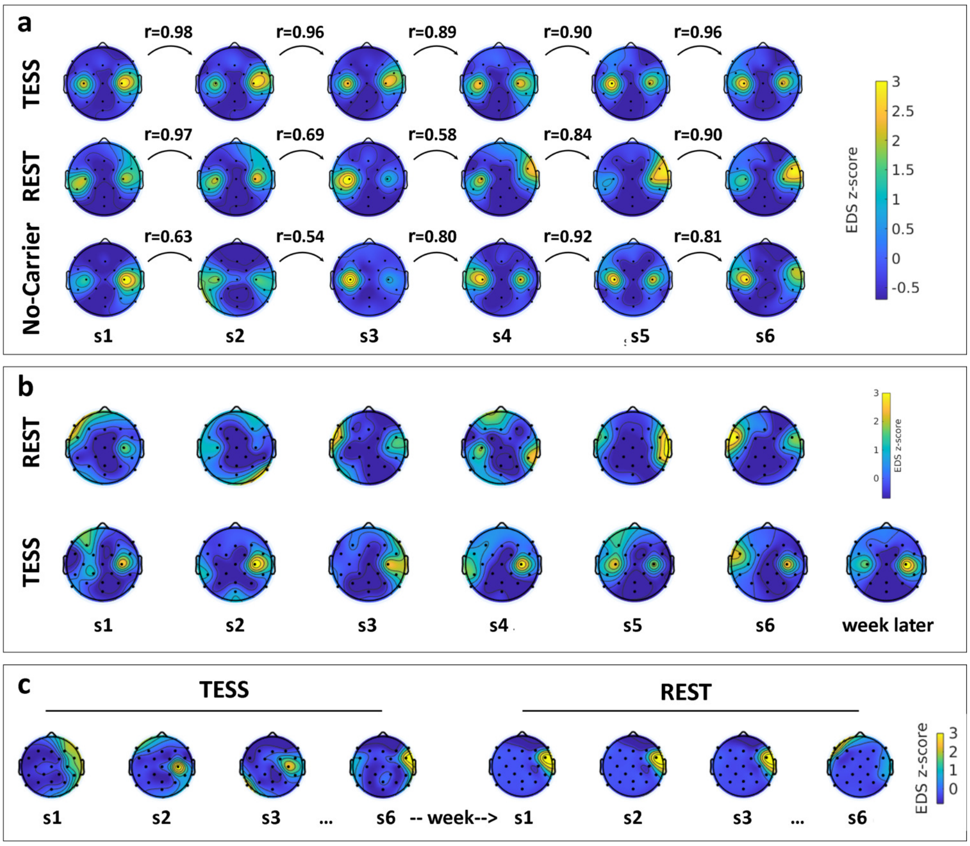


Supplementary Figure 8: **Stability of electrode discriminancy score (EDS) a**: Normalized EDS for all subjects in the TESS ($N = 14$), Rest ($N = 10$), and No-Carrier ($N = 5$) groups over the sessions of the protocol in Fig. **??**b. r-values correspond to pair-wise correlations of EDS values between consecutive sessions. **b**: Normalized EDS for the slow learners of the Cross-Over group ($N = 4$) who did TESS after 6 months of washout from Rest and with a one-week follow-up session. **c**: Normalized EDS for Patient-I during the TESS sessions and the one-week later Rest sessions.

### Supplementary Material: Transcutaneous Electrical Spinal Cord Stimulation Promotes Focal Sensorimotor Activation that Accelerates Brain-Computer Interface Skill Learning

**June 10, 2024**

#### CONSORT Reporting

The study protocol is part of the clinical trial published on ClinicalTrials.gov (NCT05183152). The original protocol includes explorations of the efficacy of different electrical stimulation techniques in enhancing the BCI training process, and it had been updated in retrospect given the incremental progression of this study that required the addition of several comparative controls to complement original findings of the core group in this study. The CONSORT enrollment flow-diagram is provided in Supplementary Fig. 9.

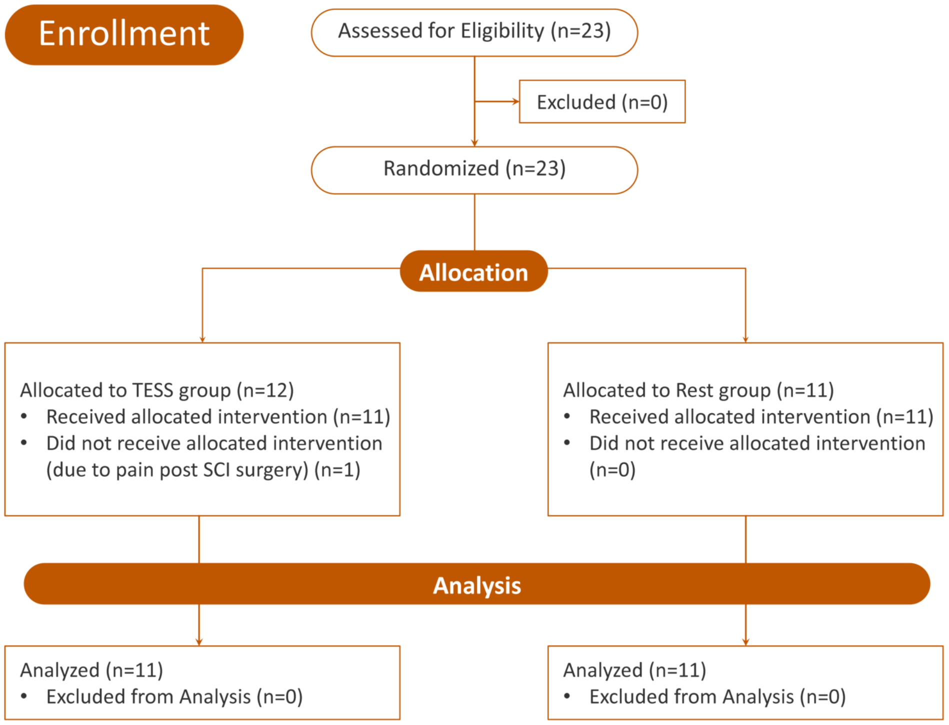


Supplementary Figure 9: **CONSORT flow diagram** . Study enrollment diagram. 23 subjects were screened and all of them were eligible and agreed to participate. They include 20 healthy participants assigned to 3 groups: 10 participants in TESS group, 10 participants in Rest group. Participants also included three patients with tetraplegia following spinal cord injury (SCI) were randomly assigned to TESS (n=2) or Rest (n=1). One patient in TESS was excluded after allocation because she was experiencing pain after her SCI surgery and had to discontinue her participation. The other two patients were re-allocated to the opposite group one week after finishing the intervention in the first group. In addition, participants in the Rest group who could not achieve BCI control were re-enrolled in the TESS group six months after their initial participation. The study was later complemented with a comparative control group of 5 participants (No-Carrier group) who had a variant of TESS without the carrier frequency.
